# Disentangling sex differences in the shared genetic architecture of posttraumatic stress disorder, traumatic experiences, and social support with body size and composition

**DOI:** 10.1101/2021.01.25.21249961

**Authors:** Carolina Muniz Carvalho, Frank R. Wendt, Gita A. Pathak, Adam X. Maihofer, Dan J. Stein, Jennifer A. Sumner, Sian M. J. Hemmings, Caroline M. Nievergelt, Karestan C. Koenen, Joel Gelernter, Sintia I. Belangero, Renato Polimanti

**Author notes:** Corresponding author: Renato Polimanti, PhD. Yale University School of Medicine, Department of Psychiatry. VA CT 116A2, 950 Campbell Avenue, West Haven, CT 06516, USA.

## Abstract

There is a well-known association of posttraumatic stress disorder (PTSD) and traumatic experiences with body size and composition, including consistent differences between sexes. However, the biology underlying these associations is unclear. To understand this complex relationship, we investigated large-scale datasets from the Psychiatric Genomic Consortium (12 823 cases and 35 648 controls), the UK Biobank (up to 360 000 individuals), and the GIANT (Genetic Investigation of Anthropometric Traits) Consortium (up to 339 224 individuals). We used genome-wide association statistics to estimate sex-specific genetic correlations (*r*_*g*_) among PTSD, traumatic experiences, social support, and multiple anthropometric traits. After multiple testing corrections (false discovery rate, FDR q<0.05), we observed 58 significant *r*_*g*_ relationships in females (e.g., childhood physical abuse and body mass index, BMI *r*_*g*_=0.245, p=3.88×10^−10^) and 21 significant *r*_*g*_ relationships in males (e.g., been involved in combat or exposed to warzone and leg fat percentage; *r*_*g*_=0.405, p=4.42×10^−10^). We performed causal inference analyses of these genetic overlaps using Mendelian randomization and latent causal variable approaches. Multiple female-specific putative causal relationships were observed linking body composition/size with PTSD (e.g., leg fat percentage➔PTSD; beta=0.319, p=3.13×10^−9^), traumatic experiences (e.g., childhood physical abuse➔waist circumference; beta=0.055, p=5.07×10^−4^), and childhood neglect (e.g., “someone to take you to doctor when needed as a child”➔BMI; beta=-0.594, p=1.09×10^−5^). In males, we observed putative causal effects linking anthropometric-trait genetic liabilities to traumatic experiences (e.g., BMI➔childhood physical abuse; beta=0.028, p=8.19×10^−3^). In conclusion, our findings provide insights regarding sex-specific causal networks linking anthropometric traits to PTSD, traumatic experiences, and social support.

## Introduction

Traumatic events include any experiences that are deeply distressing or disturbing (e.g., exposure to or threatened with death, actual or threatened serious injury, and/or sexual violence) [1]. Trauma exposure has a strong impact on mental health. Depending on trauma type and pre-trauma risk factors, a portion of trauma-exposed individuals develop posttraumatic stress disorder (PTSD) [2]. The lifetime prevalence of PTSD is higher in women (10-12%) than in men (5-6%) [3]. It is unclear whether this difference in prevalence reflects a differential vulnerability for the disorder per se, or whether because women are more likely to be exposed to events with a high conditional risk for PTSD [4]. PTSD heritability in females is two to three times higher than observed in males [5–7]. Sex-specific differences in vulnerability to PTSD may be related to mechanisms such as early stressful experiences and changes in stress-response systems (e.g., hypothalamus-pituitary-adrenal axis and sympathetic nervous system) [8– 10]. Variability across anthropometric traits reflects a highly heritable and sex-specific pre-trauma risk factor that may also be involved in the sex differences observed in trauma response and PTSD risk [11].

Anthropometric measures reflecting body composition and, in particular, fat accumulation (e.g., body mass index, BMI; waist circumference, WC; waist-hip ratio, WHR) are known to be associated with psychological stress and certain psychiatric disorders and behavioral traits [12, 13]. Anthropometric measures related to body development and growth (e.g., height) are strongly affected by early traumatic events [14, 15]. Since both PTSD and anthropometric traits show major sex differences, understanding the complex network linking PTSD and anthropometric traits could contribute to disentangling the mechanisms responsible for sex differences in the vulnerability to PTSD. In this study, our primary aim was to estimate sex-specific genetic overlap and to infer putative causal associations of anthropometric traits with PTSD and traits related to trauma and social support.

## Methods

### Study Design

This study was designed to estimate differences between sexes for genetic overlap and putative causal influences linking anthropometric measures to traits related to traumatic experiences, social support, and PTSD using large-scale genome-wide data derived from multiple cohorts: the UK Biobank (UKB), Genetic Investigation of Anthropometric Traits (GIANT), and the Psychiatric Genomics Consortium for PTSD (PGC-PTSD).

We implemented a multi-step analytic design. Firstly, we applied linkage disequilibrium score regression (LDSC) to estimate SNP-heritability and genetic correlation (*r*_*g*_) among the phenotypes of interest. For trait pairs with significant *r*_*g*_, we used the latent causal variables (LCV) method to investigate whether the *r*_*g*_ between two traits is mediated by a latent variable with a causal effect on one of the traits tested [16]. We used LCV results to select trait pairs to be tested with bidirectional polygenic risk scores (PRS). Best-fit PRS was used to determine the genetic instruments to employ for Mendelian randomization (MR) tests. Lastly, we applied MR analyses to investigate the causal relationships using genetic instruments based on a PRS analysis (Supplemental Figure 1).

**Figure 1:**
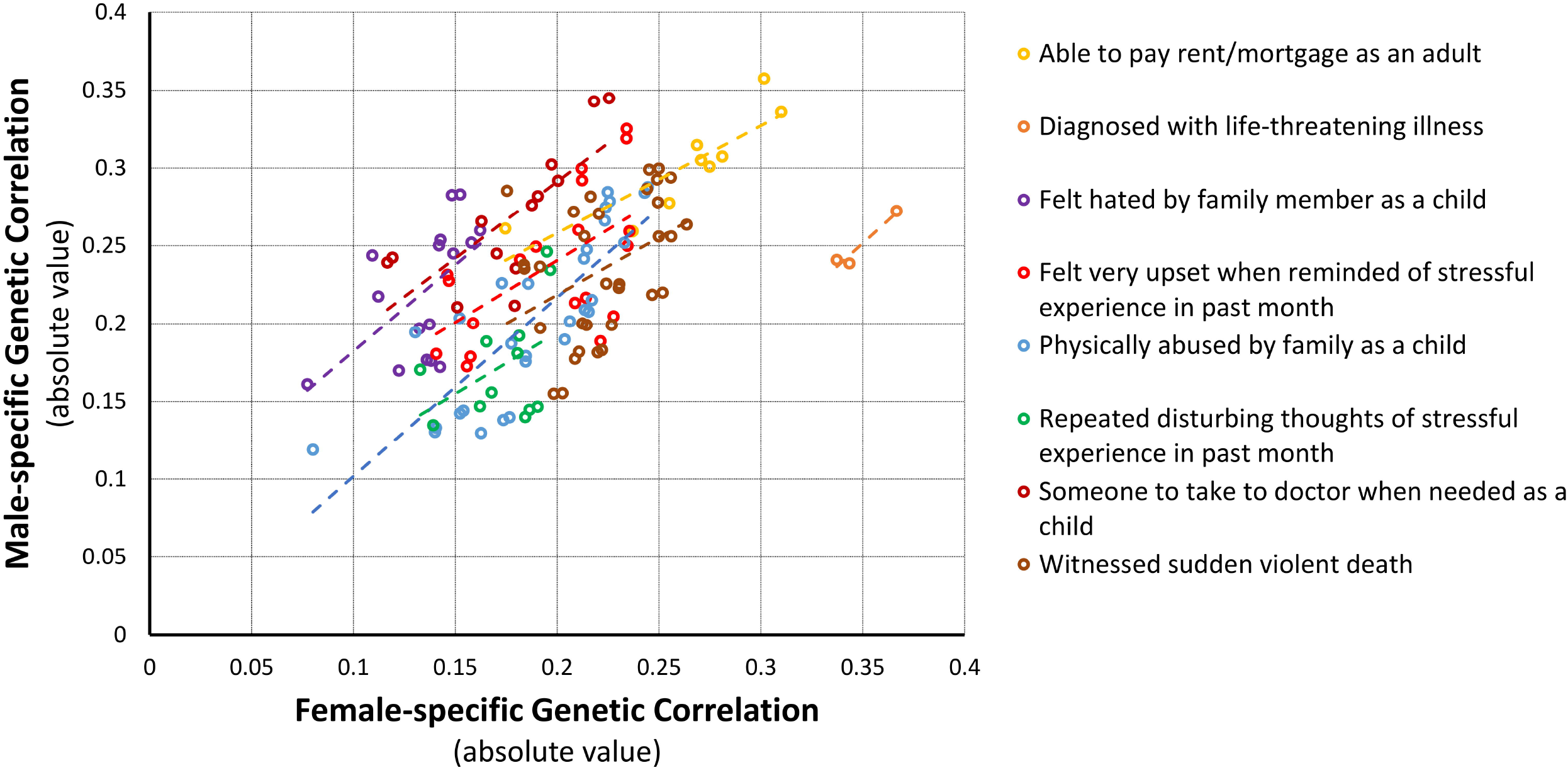
Sex-stratified genetic correlations of traits related to traumatic events and anthropometric traits surviving FDR multiple testing correction in both sexes. Linear regression (dashed line) is included in relation to the genetic correlation of each traumatic-event trait with the anthropometric traits reported in the figure.

### Data Sources

#### PTSD

We used the genome-wide association statistics from a recent GWAS of PTSD conducted by the PGC-PTSD workgroup [17]. Considering data generated from participants of European descent, we investigated data freeze 1.5 which includes subjects from PGC cohorts (12 823 cases and 35 648 controls; 40% female), but excludes the UKB [17]. We decided to not use the larger PGC-PTSD freeze 2 that includes PGC cohorts plus the UKB to avoid biases related to the sample overlap between exposure and outcome traits in the PRS and MR analyses.

#### Anthropometrics traits

We used GWAS datasets from GIANT and UKB. GIANT is an international consortium investigating genetic variants associated with human body size and shape measures, such as BMI, height, and traits related to WC (see https://portals.broadinstitute.org/collaboration/giant/index.php/Main_Page). We used sex-stratified European-ancestry genetic data from up to 171 977 females and 152 893 males [18, 19]. UKB is an international health resource that provides genetic information and data about a wide range of diseases, lifestyle habits, and behaviors in up to 502 543 individuals (aged 40–69 years) [20, 21]. Details regarding GWAS quality control criteria and methods are reported in the supplemental methods. We investigated sex-stratified GWAS association statistics of anthropometric traits derived from up to 193 000 female and 166 440 male UKB participants of European descent as defined by principal component analysis of the genetic data. The UKB traits investigated (Supplemental Table 1) are included in two data categories: “body size measures” and “impedance measures”. The “body size measures” category (UKB Field ID: 100010) includes data on body composition measures that were taken manually (i.e., standing/sitting height, waist/hip circumference, weight, and body mass index). The “impedance measures” category (UKB Field ID: 100009) include data on whole-body bio-impedance measures obtained by the Tanita BC418MA body composition analyzer (i.e., basal metabolic rate, body type, weight, body fat mass, body fat percentage, fat-free mass, water mass, predicted muscle mass and impedance for the right arm, right leg, left arm, left leg and trunk). Information regarding GIANT and UKB GWAS of anthropometric traits is reported in Supplemental Table 1.

#### Traits related to trauma and social support

We examined sex-stratified GWAS data derived from the UKB traits included in the category “traumatic events” (UKB Field ID: 145; Supplemental Table 1). These traits were assessed in up to 65 000 female participants and 51 000 male participants of European descent. These traits were assessed using the previously validated UKB online mental health questionnaire [22].

### Linkage disequilibrium score regression

Linkage Disequilibrium Score Regression (LDSC) [23, 24] (available at https://github.com/bulik/ldsc) was used to calculate the SNP-heritability estimates and *r*_*g*_ using GWAS association statistics. SNP-heritability corresponds to the proportion of phenotypic variance captured by the associated SNPs in the GWAS [25]. The *r*_*g*_ is a measure of the genetic components shared by two phenotypes investigated [26]. We investigated the SNP-heritability for anthropometric traits (38 traits from UKB and 7 traits from GIANT consortium), PTSD (PGC Freeze 1.5), and 21 traits related to traumatic experiences and social support. As recommended by LDSC developers [23, 24], we conducted the genetic correlation analysis only with the traits with heritability z-score>4 [23, 24]. A multiple testing false discovery rate correction (FDR, q<0.05) was applied to determine the statistical significance of the *r*_*g*_ calculated.

### Latent causal variable (LCV)

The latent causal variable (LCV) method postulates that the *r*_*g*_ between two traits can be mediated by a latent variable that has a causal effect on the traits tested [16]. LCV estimates the genetic causality proportion (GCP) using z-score converted per-variant effects and regression weights for genome-wide association statistics [16]. We performed an initial causal inference analysis for significant *r*_*g*_ identified via LDSC. This method differs from MR in that it uses genome-wide information rather than a select subset of the most powerful available genetic instruments. A multiple testing FDR correction (q<0.05) was applied to all GCP estimates.

LCV tests the null hypothesis that GCP=0 (i.e., there is no latent causal effect between traits). P-values and z-scores indicate the significance of the test where z-scores >> 0 indicate GCP≠ 0. GCP estimates range from −1 to 1. The magnitude of GCP indicates the proportion of causality between two traits. As recommended by O’Connor and Price, 0.7 < |GCP| < 1 indicates a fully causal relationship between the traits while |GCP| < 0.70 indicates a partial causal relationship [16]. The sign of each GCP estimate indicates the causal direction inferred from each test with GCP>0 indicating that trait 1 causes trait 2 and GCP< 0 indicating that trait 2 causes trait 1. GCP estimates were interpreted if the h^2^ z-score for both traits in a pair were >7. In Results section, we describe all LCV results in their interpreted causal direction reporting only positive GCP estimates. All LCV data and their interpretation are reported as supplementary material.

### Polygenic risk score (PRS)

Based on the significant *r*_*g*_ and LCV results, PRS was computed to define the genetic instruments for MR tests using the gtx R package embedded in PRSice v1.25 software. PRS calculates an approximate estimate of the variance explained from a multivariate regression model, and its results can identify evidence for shared genetic etiology between trait pairs. PRS was calculated using the following parameters: clumping with an LD cutoff of R^2^=0.001 in 10 000-kb windows excluding the major histocompatibility complex region of the genome because of its complex LD structure. European samples from the 1000 Genomes Project were used as the LD reference panel. To avoid overlapping individuals, we compared the GWAS datasets available depending on the cohorts: i) anthropometric traits from GIANT *vs. UKB* traits related to traumatic experiences and social support; ii) anthropometric traits from UKB and GIANT *vs*. PGC-PTSD Freeze 1.5. We applied FDR multiple testing corrections to correct the p-value for the number of PRS p-value thresholds tested, considering q<0.05 as the significance threshold.

### Mendelian randomization (MR)

MR uses genetic variants to infer causal relationships between an exposure and an outcome of interest [27]. MR relies on genetic variants satisfying three assumptions: (i) genetic variants are associated with an exposure in a specific way, (ii) the genetic variants are not associated with confounding factors linking exposure and outcome, and (iii) the genetic variants are associated with the outcome only through their association with the exposure [28]. Using the R package TwoSampleMR, we used different MR methods: random-effect inverse variance weighted (IVW) [29], MR-Egger [30], weighted median [31], simple mode [31], and weighted mode [31]. These different approaches have different sensitivities with respect to different causal scenarios that may be presented. We also conducted MR sensitivity tests to detect the presence of confounding horizontal pleiotropic effects in genetic variants (MR-Egger regression intercept and MR-PRESSO (Pleiotropy RESidual Sum and Outlier) global test) [32] and to investigate the heterogeneity of variants (IVW heterogeneity test) [32]. A leave-one-out analysis was also conducted to identify potential outliers among the variants. FDR multiple testing correction was applied to account for the number of MR tests performed. The MR analysis was conducted in accordance with the STROBE (STrenghtening the Reporting of Observational studies in Epidemiology) - MR reporting guidelines [33].

## Results

### SNP-Heritability and Genetic Correlation

The SNP-heritability was calculated for PTSD, anthropometric traits, and outcomes related to traumatic experiences and social support for both sexes, observing four traits with significant sex differences (FDR q<0.05; Supplemental Table 2). Female SNP-heritability of PTSD was much higher than that observed in males, 15.4% *vs*. 1.03% (p_sex-difference_=1.43×10^−3^). Among anthropometric traits, waist circumference adjusted for BMI (WCadjBMI) showed higher heritability in males than in females (17.7% *vs*. 11.4%, p_sex-difference_=4.91×10^−5^). Among traumatic experiences, physical violence and belittlement by partner or ex-partner as an adult showed higher SNP-heritability in females than in males (UKB Field ID 20523: 4.9% *vs*. 0.7%, p_sex-difference_=3.21×10^−4^; UKB Field ID 20521: 6.5% *vs*. 2.5%, p_sex-difference_=9.72×10^−4^). Conversely, no sex differences were observed with respect to the SNP-heritability of traits related to social support (Supplemental Table 2). In the sex-stratified *r*_*g*_ analysis, the number of significant observations was more than twice significant *r*_*g*_ in the female analysis than in the male analysis (N_females_=472 *vs*. N_males_=189; Figure 1, Supplemental Table 3). Considering the top *r*_*g*_ results for traumatic experiences, the female-specific analysis identified highly significant genetic overlap between childhood traumatic experiences (i.e., UKB Field ID: 20488 – “physically abused by a family member as a child”) and BMI (UKB Field ID: 21001; *rg*=0.245, p=3.88×10^−10^) while war-related trauma (i.e., UKB Field ID: 20527 – “been involved in combat or exposed to warzone”) was genetically correlated with peripheral fat distribution (UKB Field ID: 23111 - leg fat percentage; *r*_*g*_=0.405, p=4.42×10^−10^) in males. With respect to social support, both female and male analyses showed genetic correlation with peripheral fat distribution as the strongest result (female – UKB Field ID: 20525 – “able to pay rent/mortgage as an adult” *vs*. UKB Field ID: 23111 - leg fat percentage, *r*_*g*_=-0.31, p=3.95×10^−7^; male – UKB Field ID: 20491 – “Someone to take to doctor when needed as a child” *vs*. UKB Field ID: 23111 - leg fat percentage, *r*_*g*_=-0.34, p=3×10^−4^). Considering PTSD *r*_*g*_, we observed 19 genetically correlated traits in females while no result survived multiple testing correction in males. Similarly, to what was observed with respect to traumatic experiences and social support, the strongest finding was related to peripheral fat distribution (UKB Field ID: 23111 - Leg fat percentage; *r*_*g*_=0.224, p=4.74×10^−5^ (Figure 1, Supplemental Table 3).

### Latent Causal Variable Analysis

To test whether the significant *rg* results were due to causal effects rather than shared genetic mechanisms, we applied the LCV method and observed that 39 (females) and 16 (males) combinations tested present significant putative causal effects (FDR q<5%; Figure 2, Supplemental Table 4).

**Figure 2:**
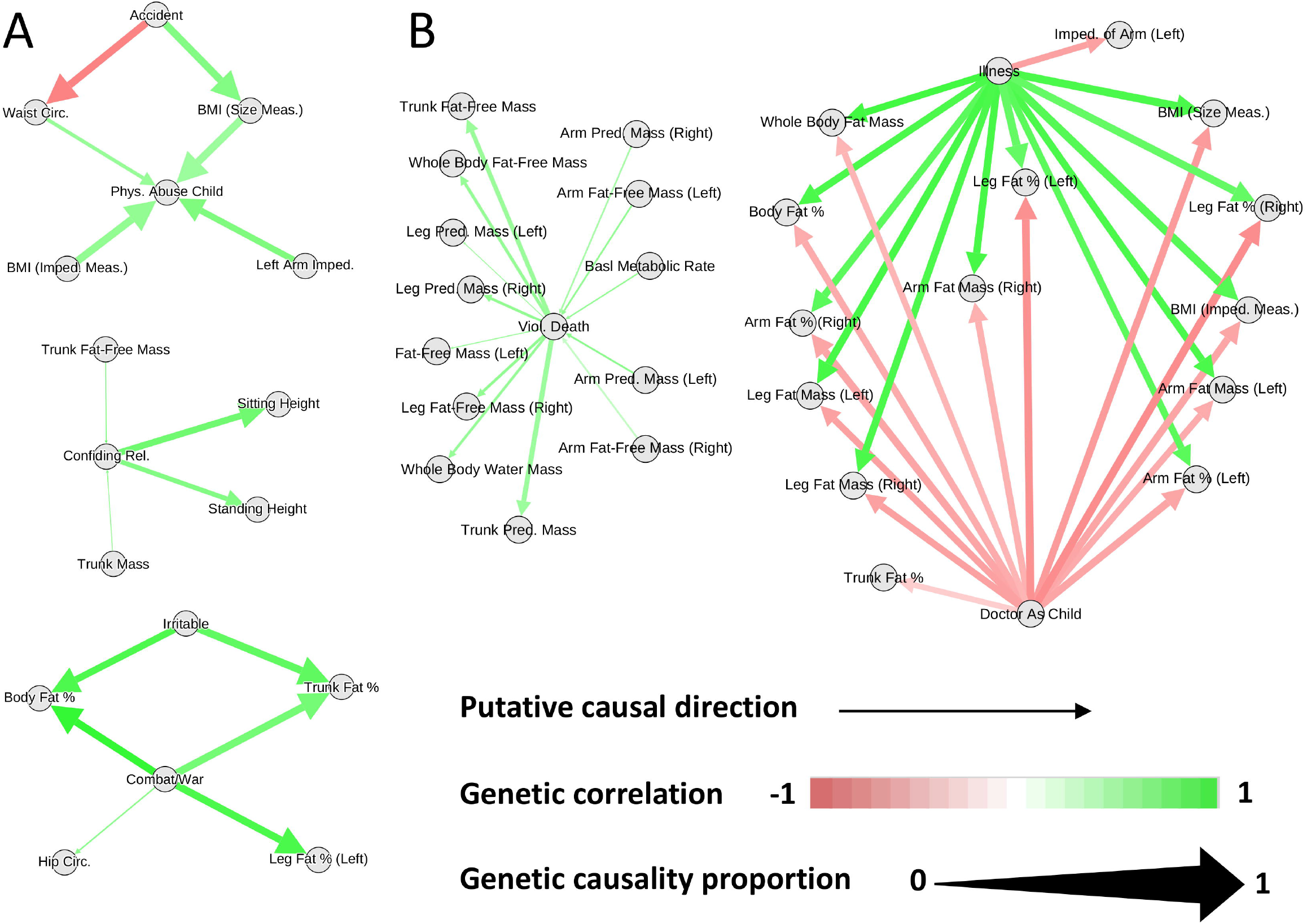
Latent causal variable network among PTSD, traumatic experiences, social support, and anthropometric traits surviving multiple testing correction. Panels A and B show male and female findings, respectively.

In females, we found that two trauma traits have large causal effects on traits linked to body composition with the strongest result being “Witnessed sudden violent death” (UKB field ID: 20530)➔ Leg predicted mass (UKB field ID: 23114, GCP=0.23, p=3.45×10^−47^). Also, the genetic liability to a social support trait (UKB field ID: 20491 – “Someone to take to doctor when needed as a child”) showed a putative effect on peripheral fat distribution (UKB field ID: 23111 - Leg fat percentage; GCP=0.76, p=1.72×10^−8^). In addition, the genetic liability to certain anthropometric traits (Supplemental Table 4) showed an association with a small but significant effect to witnessing sudden violent death (top-result: UKB field ID 23105 “Basal metabolic rate”➔ UKB field ID 20530 “Witnessed sudden violent death”; GCP=0.094, p=6.45×10^−12^).

Differently from females where multiple traumatic experiences showed putative causal effects on anthropometric traits, we observed that only three traits related to trauma have causal effects on body size and composition (Supplemental Table 4) with the strongest effect observed for “Been in serious accident/believed to be life-threatening” (UKB Field ID: 20526) with respect to anthropometric traits➔ waist circumference (UKB Field ID: 48; GCP=0.729, p=3.11×10^−14^). However, genetic liabilities to certain anthropometric traits (Supplemental Table 4) were associated with risk of childhood physical abuse in males (UKB field ID: 20488) with the strongest effect observed for waist circumference (UKB Field ID: 48)➔ childhood physical abuse (GCP=0.318, p=1.69×10^−4^). In relation to social support, we observed that the genetic liability to “been in a confiding relationship as an adult” (UKB field ID: 20522) had a moderate effect on standing and sitting height (UKB field ID: 50 and 20015, respectively; GCP=0.44, standing height, p=2.47×10^−20^; GCP=0.623, p=7.82×10^−20^), suggesting a possible nurture effect where parental genotypes affect height development. However, the genetic liability to trunk fat-free mass and trunk predicted mass (UKB field IDs: 23129 and 23130, respectively) also showed small but significant effects on been in a confiding relationship as an adult (gcp.pm=0.011, p=1.33×10^−6^; gcp.pm=0.015, p=1.17×10^−4^; respectively).

### Mendelian randomization (MR)

Similar to the strategy used in previous studies [34–37], we used PRS results to define genetic instruments for MR causal inference among the traits that showed significant genetic correlations and LCV. Accounting for the number of PRS thresholds tested, we identified 42 and six genetic instruments (i.e., sets of SNPs) significant after FDR multiple testing correction (q<0.05) in females and males, respectively (Supplemental Table 5). These were applied with respect to different MR approaches and further tested for potential biases using appropriate sensitivity analyses. We applied FDR multiple testing correction (q<0.05) to account for the number of MR tests performed. Figure 3 and Supplemental Table 6 describe the significant MR findings.

**Figure 3:**
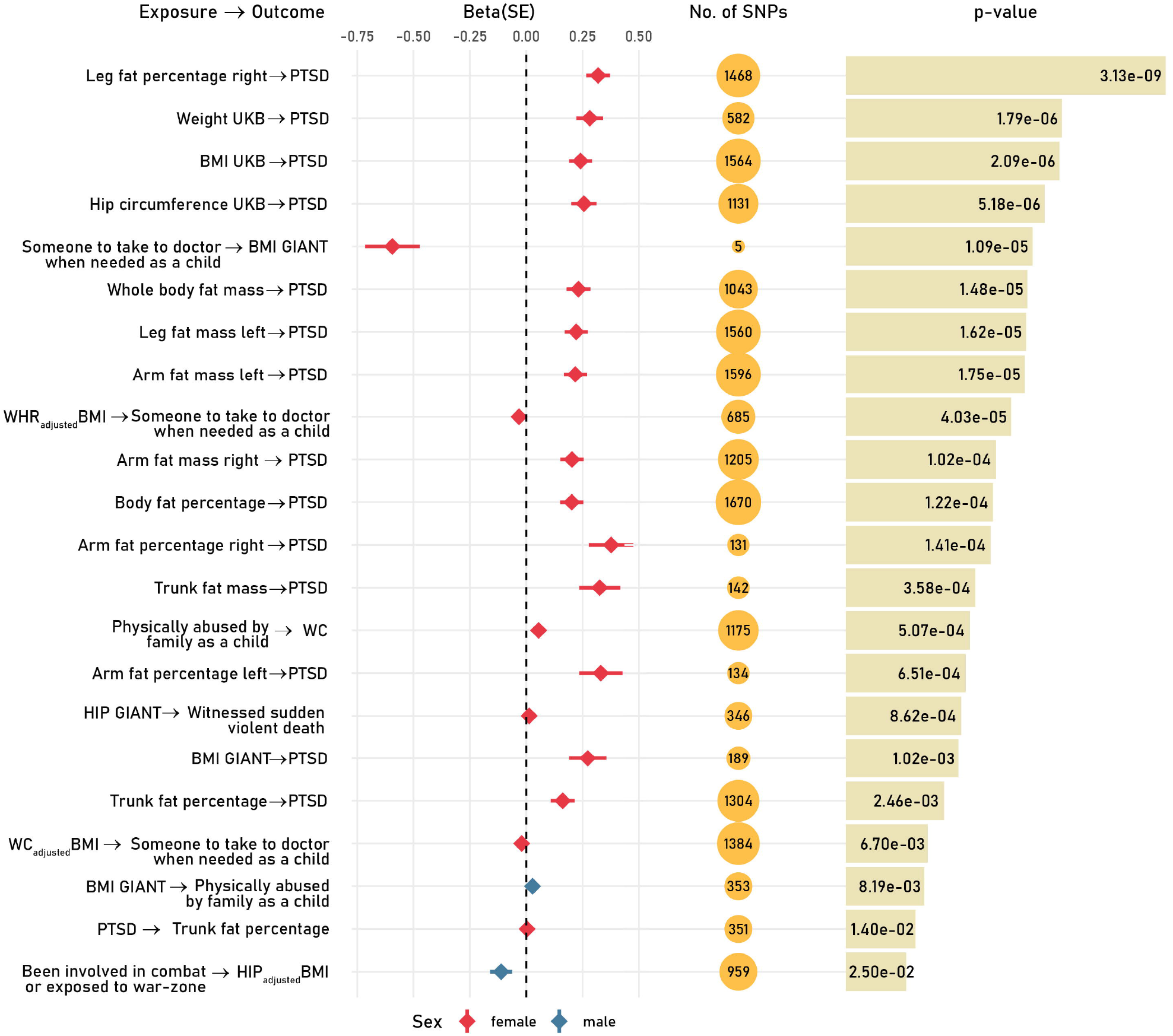
Significant Mendelian randomization (MR) tests based on the inverse variance weighted method (IVW; FDR q< 0.05). Effect size (beta) and 95% confidence interval are reported for each MR test.

In the female-specific analysis, we identified evidence of putative causal effects of multiple traits related to body fat distribution on PTSD. The strongest effect on PTSD was observed for peripheral fat distribution (UKB Field ID: 23111 – Leg fat percentage; IVW beta=0.319, p=3.13×10^−9^). We also observed a nominally-significant reverse effect of PTSD on “trunk fat percentage” (IVW beta=0.0042, p=0.014). With respect to traumatic experiences, childhood physical abuse (UKB Field ID: 20488) has a potential causal effect on WC (IVW beta=0.055, p=5.07×10^−4^). Conversely, the genetic liability to hip circumference was associated with a higher risk of having witnessed sudden violent death (UKB Field ID: 20530; IVW beta=0.0134, p=8.62×10^−4^). Additionally, the genetic liability to body shape traits (WC and BMI-adjusted WHR) showed a negative effect on childhood social support (UKB Field ID: 20491 “Someone to take to doctor when needed as a child”, WCadjBMI - IVW beta=-0.020, p=6.7×10^−3^; WHRadjBMI - IVW beta=-0.032, p=4.03×10^−5^). Conversely, childhood social support (UKB Field ID: 20491) is associated with reduced BMI (IVW beta=-0.594, p=1.09×10^−5^).

In the male-specific analysis, genetic liability to BMI showed a putative effect on childhood physical abuse (UKB Field ID: 20488; IVW beta=0.0287, p=8.19×10^−3^). An inverse effect direction was observed with respect to experiencing war-related events (UKB Field ID: 20527) with hip circumference adjusted for BMI (HIPadjBMI; IVW beta=-0.11, p=2.5×10^−2^).

## Discussion

PTSD is a multifactorial condition affecting both mental and physical health [38]. PTSD and traumatic experiences have been associated with higher BMI and obesity [39–41] with sex differences in the specific relationships among PTSD, body composition, and other domains of human health [7, 42]. Clinical studies reported that women with PTSD and a history of childhood trauma have increased BMI, adiposity, and WHR. [7, 15, 43–47]. In the present study, we used large-scale genomic datasets to understand the sex-specific associations linking PTSD, traumatic experiences, and social support with body size and composition. Our analyses highlighted that there are many more genetic correlations and putative causal effects among these traits in females than in men. This may be due to the higher vulnerability to PTSD in women than in men [3, 48] and/or to the higher heritability of PTSD in women than in men [49–53]. Additionally, similarly to PTSD-related traits, body size and composition displayed more pronounced genetic effects in women than in men [54]. With respect to the effect direction linking PTSD-related traits to body size and composition, we found a complex network of putative causal effects. In some cases, PTSD-related traits showed putative causal effects on body size and composition, while in other circumstances the effects were reversed. This suggests that the sex differences observed are not due to a single widespread mechanism linking PTSD, traumatic experiences, and social support with body size and composition. Conversely, we hypothesize that the putative causal effects observed reflect a diverse array of overlapping mechanisms linking PTSD-related traits to multiple biological systems reflected by the individual-variability of body size and composition between sexes. Indeed, anthropometric traits can reflect multiple pathways related to steroid hormone regulation, adipogenesis, lipid storage, muscle metabolism, composition, and contractile speed, skeletal growth and maturation, and lipolysis [54, 55]. Under this scenario, the effects of PTSD, traumatic experiences, and social support on body size and composition may be due to the pathogenic changes induced on the regulation of these biological pathways. However, we also observed instances where genetic liabilities to certain anthropometric traits are associated with an increased risk of PTSD, traumatic experiences, and low social support. With respect to PTSD, genetic associations with peripheral fat distribution indicate possible causal effects on PTSD risk in females. This putative effect direction appears to be female-specific (i.e., no effect was observed in males). Conversely, both female and male analyses showed the genetic liability to certain anthropometric traits to be associated with the risk of traumatic experiences. In particular, we observed putative causal effects on childhood physical abuse in both sexes. This reverse direction may reflect the complex relationship of anthropometric traits with other factors, including socioeconomic status [56], parental effects on childhood environment [57], and propensity to report traumatic events [58]. Previous studies showed that socioeconomic status is a key mediator in the association of PTSD-related traits with inflammatory biomarkers and behavioral traits [35, 59]. A recent brain-wide analysis showed that socioeconomic status has a pervasive effect on the genetics of psychiatric disorders, behavioral traits, and brain imaging phenotypes [60]. Based on these previous findings, the putative causal effect of anthropometric traits on PTSD and traumatic experience observed in our study may be affected by the pervasive association of socioeconomic status on the genetics of traits investigated. As mentioned, other factors could also play an important role. For example, the relationship could also reflect that in certain cultures people with high BMI may be more likely to be bullied or victimized and therefore traumatized.

Our results contribute to a better understanding of sex-specific mechanisms linking PTSD to the inter-individual variability of body size and composition, but our study has three main limitations. Our analyses were based on GWAS datasets generated from individuals of European descent. Since there are well-known anthropometric differences across human populations due to genetic and environmental factors [61, 62], the results described here cannot be used to make hypotheses related to non-European populations. The high PTSD polygenicity likely affected the statistical power of our causal inference analysis due to the small average per-variant effect. Finally, although we used multiple causal inference methods to account for different pleiotropy scenarios, there may be still unaccounted biases that may affect our results.

In conclusion, our study represents the largest genetic investigation of the sex-specific relationships among posttraumatic stress disorder, traumatic experiences, and social support with respect to body size and composition. These findings suggest a complex network of associations where PTSD-related traits are affected or affect anthropometric traits due to their links with multiple biological pathways. These include both shared genetic mechanisms and putative causal effects that appear more pronounced in women than in men. This suggests that both biological mechanisms and pre-trauma risk factors contribute to the higher vulnerability to PTSD observed in women.

## Data Availability

All data used to make conclusions discussed in this study are provided as Supplementary Material.

## Acknowledgments

This research was supported by the National Institutes of Health (R21 DC018098, R21 DA047527, F32 MH122058, and R01MH106595). CMC and SIB were supported by Fundação de Amparo à Pesquisa do Estado de São Paulo (FAPESP 2018/05995-4) international fellowship.

