## Supplemental Material for "Disentangling sex differences in the shared genetic architecture of posttraumatic stress disorder, traumatic experiences, and social support with body size and composition"

*UK Biobank GWAS association statistics*.

Details regarding quality control criteria and methods of this previous analysis are available at <https://github.com/Nealelab/UK_Biobank_GWAS/tree/master/imputed-v2-gwas>. Briefly, the association analyses for all phenotypes were conducted using regression models available in Hail (available at <https://github.com/hail-is/hail>) including the first 20 ancestry principal components, sex, age, age^2^, sex×age, and sex×age^2^ as covariates [1].

*GiANT association statistics*

Briefly, the sex-specific association summary statistics were corrected for residual population structure using the genomic control inflation factor (median λGC=1.01, range=0.99 – 1.08). Furthermore, SNPs were removed if minor allele count ≤ 3, deviation from Hardy-Weinberg equilibrium exact p<10^−6^, SNP call rate<95%, or low imputation quality (below 0.3 for MACH, 0.4 for IMPUTE, and 0.8 for PLINK) [2, 3].

*PGC PTSD* *association statistics*

Briefly, SNPs were removed if minor allele frequency < 5%, deviation from Hardy-Weinberg equilibrium p<10^−6^, SNP call rate<98%, and a > 2% difference in missing genotypes between cases and control. The association sex-stratified analyses for studies with unrelated subjects, additive model via logistic regression in PLINK 1.9 were used to test the association with PTSD, including the first five PC’s as covariates; and in the UKB cohort, the association studies the six PC’s in the linear regression [4].

**Supplemental Figure**

**Figure 1:** Flow chart of study design

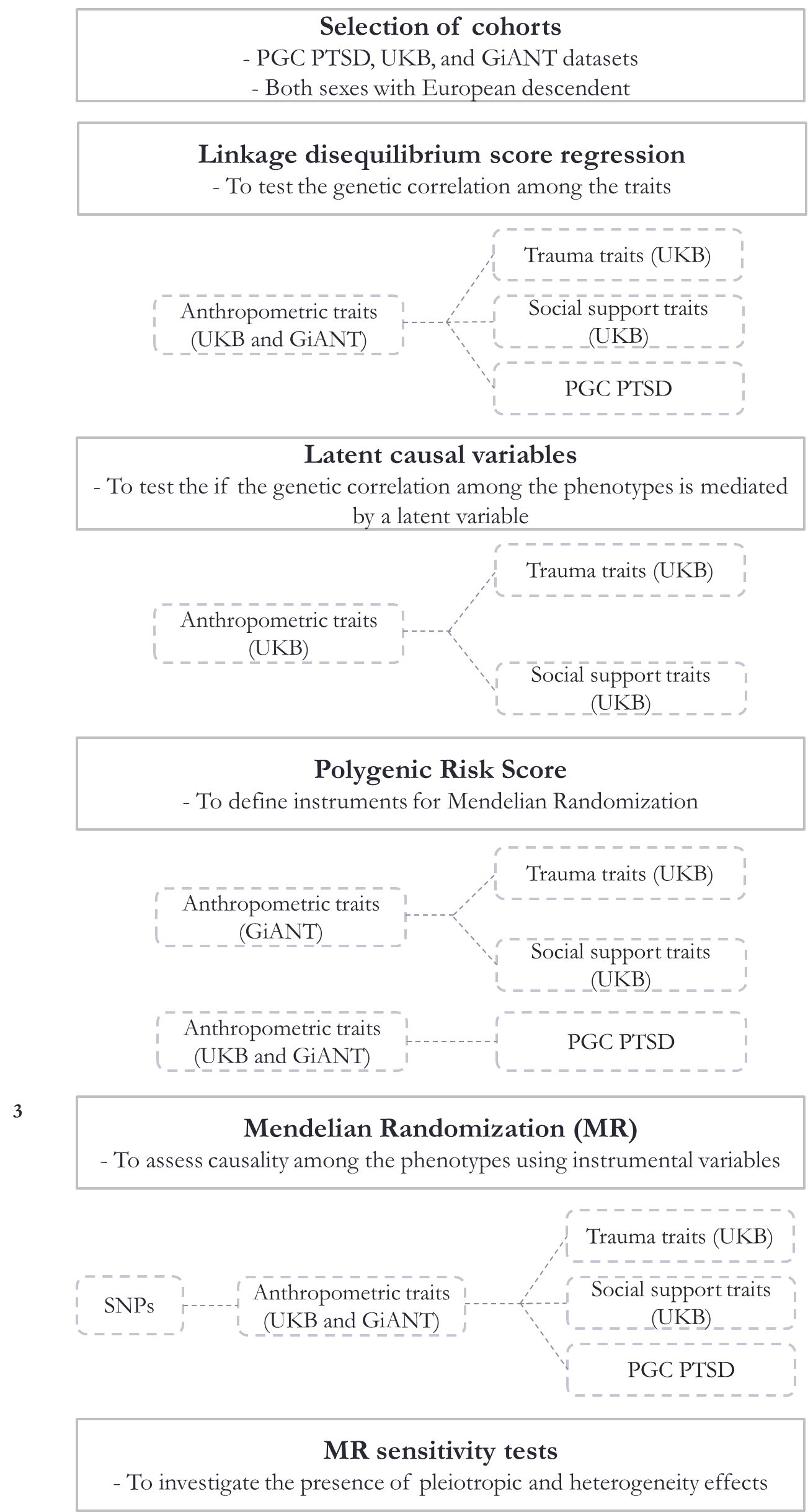

PGC PTSD: Psychiatric Genomics Consortium Posttraumatic stress disorder. UKB: UK Biobank. GiANT: Genetic Investigation of ANthropometric Traits.

**Supplemental Tables**

**Supplemental Table 1:** Phenotypic traits investigated in this study. Sex-stratified sample size is reported for each trait (F: females; M: Male).

| **Psychiatric Genomics Consortium** | **Abbreviation** | **Sex- stratified Sample size** |
| --- | --- | --- |
| Posttraumatic Stress Disorder | PTSD | F_cases/controls_=6 128/9 528  M_cases/controls_=6 364/23 905 |
| **UK Biobank (UKB)** | **UKB Field ID** | **Sex- stratified Sample size** |
| **Traumatic events (UKB Category 145)** | | |
| Felt irritable or had angry outbursts in past month | 20494 | F=33 321  M=19 495 |
| Avoided activities or situations because of previous stressful experience in past month | 20495 | F=65 984  M=51 884 |
| Felt distant from other people in past month | 20496 | F=33 325  M=19 497 |
| Repeated disturbing thoughts of stressful experience in past month | 20497 | F=66 006  M=51 894 |
| Felt very upset when reminded of stressful experience in past month | 20498 | F=65 995  M=51 898 |
| Felt loved as a child | 20489 | F=65 867  M=51 757 |
| Physically abused by family as a child | 20488 | F=65 969  M=51 869 |
| Felt hated by family member as a child | 20487 | F=65 904  M=51 845 |
| Sexually molested as a child | 20490 | F=65 117  M=51 656 |
| Someone to take to doctor when needed as a child | 20491 | F=65 629  M=51 672 |
| Been in a confiding relationship as an adult | 20522 | F=64 141  M=50 958 |
| Physical violence by partner or ex-partner as an adult | 20523 | F=65 891  M=51 855 |
| Belittlement by partner or ex-partner as an adult | 20521 | F=65 906  M=51 835 |
| Sexual interference by partner or ex-partner without consent as an adult | 20524 | F=65 854  M=51 873 |
| Able to pay rent/mortgage as an adult | 20525 | F=65 093  M=51 203 |
| Victim of sexual assault | 20531 | F=65 029  M=51 642 |
| Victim of physically violent crime | 20529 | F=65 959  M=51 887 |
| Been in serious accident believed to be life-threatening | 20526 | F=66 030  M=51 892 |
| Witnessed sudden violent death | 20530 | F=65 994  M=51 868 |
| Diagnosed with life-threatening illness | 20528 | F=65 818  M=51 799 |
| Been involved in combat or exposed to war-zone | 20527 | F=66 077  M=51 877 |
| **Impedance measures (UKB Category 100009)** | | |
| **Impedance measures (UKB Category 100009)** | | |
| Weight | 23098 | F=190 993  M=163 845 |
| Body mass index (BMI) | 23104 | F=190 990  M=163 841 |
| Leg fat-free mass (right) | 23113 | F=190 972  M=163 826 |
| Leg predicted mass (left) | 23118 | F=190 955  M=163 811 |
| Leg predicted mass (right) | 23114 | F=190 972  M=163 826 |
| Arm fat percentage (left) | 23123 | F=190 921  M=163 786 |
| Arm fat percentage (right) | 23119 | F=190 952  M=163 808 |
| Arm fat mass (left) | 23124 | F=190 903  M=163 770 |
| Arm fat mass (right) | 23120 | F=190 931  M=163 805 |
| Arm fat-free mass (right) | 23121 | F=190 934  M=163 798 |
| Arm fat-free mass (left) | 23125 | F=190 904  M=163 764 |
| Arm predicted mass (left) | 23126 | F=190 897  M=163 756 |
| Arm predicted mass (right) | 23122 | F=190 930  M=163 796 |
| Trunk fat percentage | 23127 | F=190 879  M=163 740 |
| Trunk fat mass | 23128 | F=190 867  M=163 730 |
| Trunk fat-free mass | 23129 | F=190 833  M=163 697 |
| Trunk predicted mass | 23130 | F=190 816  M=163 678 |
| Basal metabolic rate | 23105 | F=190 987  M=163 838 |
| Body fat percentage | 23099 | F=190 991  M=163 637 |
| Whole body fat mass | 23100 | F=190 941  M=163 303 |
| Whole body fat-free mass | 23101 | F=190 993  M=163 815 |
| Whole body water mass | 23102 | F=190 992  M=163 842 |
| Leg fat percentage (left) | 23115 | F=190 969  M=163 822 |
| Leg fat percentage (right) | 23111 | F=190 977  M=163 834 |
| Leg fat mass (left) | 23116 | F=190 967  M=163 821 |
| Leg fat mass (right) | 23112 | F=190 975  M=163 832 |
| Leg fat-free mass (left) | 23117 | F=190 958  M=163 813 |
| Impedance of whole body | 23106 | F=190 958  M=163 837 |
| **Impedance measures (UKB Category 100009)** | | |
| Impedance of arm (left) | 23110 | F=190 974  M=163 833 |
| Impedance of arm (right) | 23109 | F=190 961  M=163 831 |
| Impedance of leg (left) | 23108 | F=190 977  M=163 834 |
| Impedance of leg (right) | 23107 | F=190 980  M=163 837 |
| **Body sizes measures (UKB Category 100010)** | | |
| Waist circumference | 48 | F=193 828  M=166 736 |
| Weight | 21002 | F=193 627  M=166 489 |
| Body mass index (BMI) | 21001 | F=193 570  M=166 413 |
| Hip circumference | 49 | F=193 814  M=166 707 |
| Standing height | 50 | F=193 785  M=166 603 |
| Sitting height | 20015 | F=193 600  M=166 466 |
| **Genetic Investigation of ANthropometric Traits (GIANT)** | **Abbreviation** | **Sex- stratified Sample size** |
| Hip circumference | HIP | F=118 528  M=94 790 |
| Waist circumference | WC | F=127 998  M=104 406 |
| Waist-to-hip ratio | WHR | F=118 004  M=94 434 |
| Body mass index | BMI | F=171 977  M=152 893 |
| Hip circumference adjusted for BMI | HIPadjBMI | F=117 340  M=93 965 |
| Waist circumference adjusted for BMI | WCadjBMI | F=127 470  M=104 079 |
| Waist-to-hip ratio adjusted for BMI | WHRadjBMI | F=116 742  M=93 480 |

**Supplemental Table 2:** Sex-stratified SNP-heritability of PTSD, anthropometric traits, and traits related to traumatic experiences and social support. The definition of the phenotype IDs is available in Supplemental Table 1. H^2^: SNP-heritability. SE: standard error of SNP-heritability. FDR: false discovery rate. BMI: body mass index. HIPadjBMI: hip circumference adjusted for BMI. WC: waist circumference. WCadjBMI: waist circumference adjusted for BMI. WHR: waist-to-hip ratio. WHRadjBMI: waist-to-hip ratio adjusted by BMI.

| **Phenotype ID** | **Males** | | **Females** | | **M v. F Difference** | | |
| --- | --- | --- | --- | --- | --- | --- | --- |
|  | **H^2^** | **SE** | **H^2^** | **SE** | **z** | **p** | **FDR q** |
| WCadjBMI | 0.1773 | 0.0125 | 0.1138 | 0.0094 | 4.0601 | 4.91E-05 | 0.00329 |
| 20523 | 0.0066 | 0.0091 | 0.0488 | 0.0074 | -3.5979 | 3.21E-04 | 0.01 |
| 20521 | 0.0248 | 0.0092 | 0.0648 | 0.0079 | -3.2986 | 9.72E-04 | 0.02171 |
| PTSD | 0.0103 | 0.0253 | 0.1541 | 0.0373 | -3.1905 | 1.42E-03 | 0.02379 |
| 20527 | 0.0343 | 0.0086 | 0.0067 | 0.0062 | 2.6033 | 9.23E-03 | 0.10050 |
| 20487 | 0.0318 | 0.009 | 0.0612 | 0.007 | -2.5786 | 9.92E-03 | 0.10050 |
| HIPadjBMI | 0.2057 | 0.0166 | 0.1541 | 0.012 | 2.5191 | 1.18E-02 | 0.10050 |
| 20524 | 0.0046 | 0.008 | 0.0318 | 0.0073 | -2.5115 | 1.20E-02 | 0.10050 |
| 20498 | 0.0369 | 0.0091 | 0.0671 | 0.0084 | -2.4386 | 1.47E-02 | 0.10943 |
| 20531 | 0.019 | 0.0083 | 0.0459 | 0.0076 | -2.3903 | 1.68E-02 | 0.11256 |
| 23130 | 0.3435 | 0.0176 | 0.2931 | 0.0123 | 2.3472 | 1.89E-02 | 0.11278 |
| 23129 | 0.3444 | 0.0176 | 0.2944 | 0.0124 | 2.3224 | 2.02E-02 | 0.11278 |
| 20530 | 0.0414 | 0.0088 | 0.0186 | 0.0065 | 2.0840 | 3.72E-02 | 0.18425 |
| 20489 | 0.0745 | 0.01 | 0.101 | 0.008 | -2.0693 | 3.85E-02 | 0.18425 |
| 20495 | 0.0202 | 0.0085 | 0.0409 | 0.0079 | -1.7838 | 7.45E-02 | 0.33277 |
| 23110 | 0.2612 | 0.011 | 0.239 | 0.0092 | 1.5481 | 1.22E-01 | 0.44348 |
| 23105 | 0.3257 | 0.015 | 0.2964 | 0.0118 | 1.5352 | 1.25E-01 | 0.44348 |
| 23126 | 0.3023 | 0.0139 | 0.2757 | 0.011 | 1.5006 | 1.33E-01 | 0.44348 |
| 23125 | 0.3031 | 0.0139 | 0.2766 | 0.011 | 1.4950 | 1.35E-01 | 0.44348 |
| WHR | 0.1405 | 0.0114 | 0.1172 | 0.0107 | 1.4903 | 1.36E-01 | 0.44348 |
| 23109 | 0.2628 | 0.0107 | 0.2418 | 0.0093 | 1.4813 | 1.39E-01 | 0.44348 |
| 23101 | 0.3346 | 0.0161 | 0.3063 | 0.0125 | 1.3884 | 1.65E-01 | 0.50250 |
| 23102 | 0.3334 | 0.016 | 0.3061 | 0.0125 | 1.3446 | 1.79E-01 | 0.52143 |
| 20491 | 0.0236 | 0.009 | 0.0398 | 0.0088 | -1.2870 | 1.98E-01 | 0.55275 |
| 20525 | 0.0167 | 0.0082 | 0.0304 | 0.0074 | -1.2403 | 2.15E-01 | 0.57620 |
| 23122 | 0.3051 | 0.0137 | 0.2839 | 0.0116 | 1.1810 | 2.38E-01 | 0.61331 |
| 23121 | 0.3054 | 0.0137 | 0.2847 | 0.0116 | 1.1531 | 2.49E-01 | 0.61789 |
| 21002 | 0.285 | 0.0115 | 0.2679 | 0.01 | 1.1221 | 2.62E-01 | 0.62693 |
| 23128 | 0.2395 | 0.0093 | 0.2529 | 0.0087 | -1.0522 | 2.93E-01 | 0.64069 |
| WC | 0.1813 | 0.0168 | 0.1611 | 0.0098 | 1.0386 | 2.99E-01 | 0.64069 |
| 23123 | 0.2358 | 0.0092 | 0.2489 | 0.0087 | -1.0346 | 3.01E-01 | 0.64069 |
| 23098 | 0.2843 | 0.0115 | 0.2687 | 0.01 | 1.0236 | 3.06E-01 | 0.64069 |
| HIP | 0.191 | 0.0151 | 0.174 | 0.0105 | 0.9243 | 3.55E-01 | 0.72076 |
| 48 | 0.2217 | 0.0091 | 0.2114 | 0.0082 | 0.8409 | 4.00E-01 | 0.78824 |
| 23106 | 0.2807 | 0.0119 | 0.2681 | 0.0103 | 0.8006 | 4.23E-01 | 0.79842 |
| 20522 | 0.0348 | 0.0088 | 0.0439 | 0.0074 | -0.7915 | 4.29E-01 | 0.79842 |
| 23119 | 0.238 | 0.0091 | 0.2468 | 0.0087 | -0.6990 | 4.85E-01 | 0.82745 |
| 21001 | 0.2668 | 0.0105 | 0.257 | 0.0095 | 0.6921 | 4.89E-01 | 0.82745 |
| WHRadjBMI | 0.1278 | 0.0111 | 0.1159 | 0.0132 | 0.6900 | 4.90E-01 | 0.82745 |
| 20488 | 0.0559 | 0.0091 | 0.064 | 0.0076 | -0.6832 | 4.94E-01 | 0.82745 |
| 20529 | 0.028 | 0.0095 | 0.0199 | 0.0082 | 0.6454 | 5.19E-01 | 0.83516 |
| 23104 | 0.2666 | 0.0106 | 0.2576 | 0.0096 | 0.6293 | 5.29E-01 | 0.83516 |
| 23107 | 0.2578 | 0.0115 | 0.2484 | 0.0099 | 0.6195 | 5.36E-01 | 0.83516 |
| 20496 | 0.0282 | 0.0237 | 0.0441 | 0.0152 | -0.5647 | 5.72E-01 | 0.87100 |
| 20490 | 0.0239 | 0.0084 | 0.03 | 0.0074 | -0.5449 | 5.86E-01 | 0.87249 |
| 23111 | 0.2385 | 0.0082 | 0.2443 | 0.0079 | -0.5094 | 6.10E-01 | 0.87969 |
| 23108 | 0.2577 | 0.0113 | 0.2505 | 0.0099 | 0.4793 | 6.32E-01 | 0.87969 |
| 23100 | 0.2446 | 0.0093 | 0.2506 | 0.0088 | -0.4686 | 6.39E-01 | 0.87969 |
| 23099 | 0.2465 | 0.009 | 0.2414 | 0.0081 | 0.4212 | 6.74E-01 | 0.87969 |
| 23124 | 0.2414 | 0.0098 | 0.2468 | 0.0091 | -0.4038 | 6.86E-01 | 0.87969 |
| 20497 | 0.0453 | 0.0092 | 0.0499 | 0.007 | -0.3979 | 6.91E-01 | 0.87969 |
| 23114 | 0.2994 | 0.0137 | 0.2923 | 0.0118 | 0.3927 | 6.95E-01 | 0.87969 |
| 23113 | 0.299 | 0.0137 | 0.2921 | 0.0118 | 0.3816 | 7.03E-01 | 0.87969 |
| 20015 | 0.3658 | 0.0229 | 0.3543 | 0.0207 | 0.3725 | 7.09E-01 | 0.87969 |
| 49 | 0.2361 | 0.0101 | 0.2316 | 0.0092 | 0.3294 | 7.42E-01 | 0.90389 |
| BMI | 0.1745 | 0.0113 | 0.1702 | 0.0089 | 0.2989 | 7.65E-01 | 0.91214 |
| 23112 | 0.2418 | 0.0089 | 0.2454 | 0.009 | -0.2844 | 7.76E-01 | 0.91214 |
| 20526 | 0.0216 | 0.0094 | 0.0239 | 0.0065 | -0.2013 | 8.41E-01 | 0.92630 |
| 23120 | 0.2441 | 0.0099 | 0.2467 | 0.0091 | -0.1934 | 8.47E-01 | 0.92630 |
| 23115 | 0.2411 | 0.008 | 0.2432 | 0.0079 | -0.1868 | 8.52E-01 | 0.92630 |
| 20494 | 0.0437 | 0.0229 | 0.0388 | 0.0137 | 0.1836 | 8.54E-01 | 0.92630 |
| 23118 | 0.2961 | 0.0133 | 0.2932 | 0.0117 | 0.1637 | 8.70E-01 | 0.92630 |
| 23117 | 0.2959 | 0.0134 | 0.293 | 0.0118 | 0.1624 | 8.71E-01 | 0.92630 |
| 23127 | 0.2357 | 0.0091 | 0.2341 | 0.0081 | 0.1313 | 8.96E-01 | 0.93800 |
| 20528 | 0.0206 | 0.0082 | 0.0215 | 0.0071 | -0.0830 | 9.34E-01 | 0.96274 |
| 23116 | 0.2445 | 0.009 | 0.2437 | 0.009 | 0.0629 | 9.50E-01 | 0.96439 |
| 50 | 0.4828 | 0.0243 | 0.4831 | 0.0242 | -0.0087 | 9.93E-01 | 0.99300 |

**Supplemental Table 3:** Sex-stratified Genetic correlations among PTSD, trauma, and social support traits vs anthropometric traits surviving FDR multiple testing correction (q<0.05). The definition of the phenotype IDs is available in Supplemental Table 1. rg: genetic correlation estimate. SE: standard error. p: p-value

| **PTSD, trauma, and social support traits** | **Anthropometric traits** | **rg** | **SE** | **p** | **FDR q** | **Sex** |
| --- | --- | --- | --- | --- | --- | --- |
| 20488 | 21001 | 0.2445 | 0.0391 | 3.88E-10 | 2.92E-07 | female |
| 20488 | 23104 | 0.243 | 0.0395 | 7.32E-10 | 2.92E-07 | female |
| 20498 | 23115 | 0.234 | 0.0399 | 4.58E-09 | 8.84E-07 | female |
| 20498 | 23111 | 0.2341 | 0.0401 | 5.47E-09 | 8.84E-07 | female |
| 20498 | 21001 | 0.2355 | 0.0406 | 6.57E-09 | 8.84E-07 | female |
| 20498 | 23104 | 0.2346 | 0.0405 | 7.24E-09 | 8.84E-07 | female |
| 20488 | 23110 | -0.233 | 0.0404 | 7.75E-09 | 8.84E-07 | female |
| 20488 | 23112 | 0.2258 | 0.0396 | 1.21E-08 | 1.21E-06 | female |
| 20523 | 23104 | 0.251 | 0.0443 | 1.51E-08 | 1.34E-06 | female |
| 20523 | 21001 | 0.2482 | 0.0442 | 1.96E-08 | 1.46E-06 | female |
| 20488 | 23120 | 0.2171 | 0.0388 | 2.17E-08 | 1.46E-06 | female |
| 20488 | 23116 | 0.2234 | 0.0399 | 2.19E-08 | 1.46E-06 | female |
| 20498 | 23119 | 0.2277 | 0.041 | 2.81E-08 | 1.59E-06 | female |
| 20488 | 23111 | 0.2249 | 0.0406 | 2.96E-08 | 1.59E-06 | female |
| 20488 | 23106 | -0.2136 | 0.0385 | 2.99E-08 | 1.59E-06 | female |
| 20488 | 23124 | 0.2155 | 0.039 | 3.26E-08 | 1.6E-06 | female |
| 20523 | 23115 | 0.2406 | 0.0436 | 3.40E-08 | 1.60E-06 | female |
| 20488 | 23115 | 0.2238 | 0.0408 | 4.22E-08 | 1.85E-06 | female |
| 20523 | 23111 | 0.2391 | 0.0437 | 4.41E-08 | 1.85E-06 | female |
| 20523 | 23119 | 0.2345 | 0.0429 | 4.67E-08 | 1.86E-06 | female |
| 20498 | 23123 | 0.2213 | 0.041 | 6.58E-08 | 2.46E-06 | female |
| 20488 | 23119 | 0.2062 | 0.0382 | 6.77E-08 | 2.46E-06 | female |
| 20523 | 23123 | 0.2274 | 0.0427 | 1.02E-07 | 3.54E-06 | female |
| 20488 | 48 | 0.2146 | 0.0407 | 1.35E-07 | 4.49E-06 | female |
| 20488 | 23109 | -0.2132 | 0.0406 | 1.48E-07 | 4.72E-06 | female |
| 20488 | 23123 | 0.2037 | 0.0388 | 1.55E-07 | 4.76E-06 | female |
| 20498 | 23120 | 0.2141 | 0.0409 | 1.65E-07 | 4.88E-06 | female |
| 20498 | 23112 | 0.212 | 0.0406 | 1.83E-07 | 5.22E-06 | female |
| 20490 | 21001 | 0.3312 | 0.0636 | 1.90E-07 | 5.23E-06 | female |
| 20498 | 23116 | 0.2122 | 0.0408 | 1.98E-07 | 5.27E-06 | female |
| 20498 | 23124 | 0.2088 | 0.0404 | 2.37E-07 | 6.10E-06 | female |
| 20490 | 23104 | 0.3259 | 0.0634 | 2.73E-07 | 6.81E-06 | female |
| 20525 | 23111 | -0.31 | 0.0611 | 3.95E-07 | 9.55E-06 | female |
| 20525 | 23115 | -0.3016 | 0.0596 | 4.18E-07 | 9.77E-06 | female |
| 20490 | 23112 | 0.3106 | 0.0615 | 4.36E-07 | 9.77E-06 | female |
| 20490 | 23116 | 0.3132 | 0.062 | 4.41E-07 | 9.77E-06 | female |
| 20523 | 23120 | 0.2229 | 0.0442 | 4.53E-07 | 9.77E-06 | female |
| 20490 | 23120 | 0.3079 | 0.0612 | 4.92E-07 | 1.03E-05 | female |
| 20523 | 23116 | 0.2224 | 0.0447 | 6.38E-07 | 1.31E-05 | female |
| 20523 | 23112 | 0.2195 | 0.0446 | 8.64E-07 | 1.72E-05 | female |
| 20523 | 23124 | 0.2165 | 0.0442 | 9.82E-07 | 1.90E-05 | female |
| 20490 | 23124 | 0.2981 | 0.061 | 1.03E-06 | 1.9E-05 | female |
| 20498 | 48 | 0.2104 | 0.0431 | 1.04E-06 | 1.90E-05 | female |
| 20488 | 21002 | 0.1847 | 0.0378 | 1.05E-06 | 1.9E-05 | female |
| 20488 | 23098 | 0.1846 | 0.038 | 1.19E-06 | 2.11E-05 | female |
| 20488 | 23100 | 0.1858 | 0.0387 | 1.58E-06 | 2.74E-05 | female |
| 20490 | 48 | 0.2913 | 0.061 | 1.77E-06 | 2.96E-05 | female |
| 20525 | 21001 | -0.2811 | 0.0589 | 1.78E-06 | 2.96E-05 | female |
| 20490 | 21002 | 0.2635 | 0.0552 | 1.85E-06 | 3.01E-05 | female |
| 20488 | 49 | 0.1775 | 0.0374 | 2.13E-06 | 3.38E-05 | female |
| 20521 | 23104 | 0.1815 | 0.0383 | 2.16E-06 | 3.38E-05 | female |
| 20490 | 23098 | 0.2591 | 0.0551 | 2.53E-06 | 3.88E-05 | female |
| 20523 | 23099 | 0.2034 | 0.0433 | 2.66E-06 | 4.01E-05 | female |
| 20521 | 21001 | 0.1793 | 0.0385 | 3.11E-06 | 4.56E-05 | female |
| 20490 | 23100 | 0.2723 | 0.0584 | 3.14E-06 | 4.56E-05 | female |
| 20498 | 23099 | 0.1896 | 0.0407 | 3.28E-06 | 4.67E-05 | female |
| 20490 | 23119 | 0.2964 | 0.0639 | 3.46E-06 | 4.84E-05 | female |
| 20490 | 23123 | 0.2952 | 0.0638 | 3.78E-06 | 5.13E-05 | female |
| 20525 | 23104 | -0.2749 | 0.0595 | 3.79E-06 | 5.13E-05 | female |
| 20490 | 23111 | 0.3034 | 0.0657 | 3.86E-06 | 5.13E-05 | female |
| 20490 | 23115 | 0.3066 | 0.0664 | 3.95E-06 | 5.17E-05 | female |
| 20521 | 23110 | -0.1885 | 0.0409 | 4.12E-06 | 5.30E-05 | female |
| 20521 | 23111 | 0.1778 | 0.0387 | 4.27E-06 | 5.41E-05 | female |
| 20488 | 23125 | 0.1767 | 0.0387 | 4.93E-06 | 6.15E-05 | female |
| 20525 | 23112 | -0.2706 | 0.0594 | 5.25E-06 | 6.32E-05 | female |
| 20525 | 23116 | -0.2687 | 0.059 | 5.29E-06 | 6.32E-05 | female |
| 20521 | 23115 | 0.1753 | 0.0385 | 5.31E-06 | 6.32E-05 | female |
| 20528 | 23100 | 0.3706 | 0.0817 | 5.67E-06 | 6.65E-05 | female |
| 20525 | 23119 | -0.2653 | 0.0588 | 6.43E-06 | 7.38E-05 | female |
| 20528 | 23128 | 0.3597 | 0.0798 | 6.57E-06 | 7.38E-05 | female |
| 20528 | 23098 | 0.3637 | 0.0807 | 6.57E-06 | 7.38E-05 | female |
| 20498 | 23100 | 0.1818 | 0.0404 | 6.66E-06 | 7.38E-05 | female |
| 20528 | 21002 | 0.3664 | 0.0815 | 6.92E-06 | 7.56E-05 | female |
| 20528 | 23120 | 0.3741 | 0.0835 | 7.42E-06 | 8.00E-05 | female |
| 20489 | 23104 | -0.1351 | 0.0303 | 8.20E-06 | 8.52E-05 | female |
| 20488 | 23099 | 0.1729 | 0.0388 | 8.29E-06 | 8.52E-05 | female |
| 20488 | 23126 | 0.1736 | 0.039 | 8.29E-06 | 8.52E-05 | female |
| 20525 | 23099 | -0.2551 | 0.0572 | 8.33E-06 | 8.52E-05 | female |
| 20525 | 23123 | -0.2619 | 0.0589 | 8.73E-06 | 8.82E-05 | female |
| 20528 | 48 | 0.3666 | 0.0829 | 9.86E-06 | 9.84E-05 | female |
| 20525 | 23120 | -0.259 | 0.0588 | 1.07E-05 | 1.05E-04 | female |
| 20528 | 23124 | 0.3636 | 0.0827 | 1.08E-05 | 1.05E-04 | female |
| 20528 | 23116 | 0.3645 | 0.083 | 1.12E-05 | 1.08E-04 | female |
| 20523 | 48 | 0.2035 | 0.0466 | 1.28E-05 | 1.20E-04 | female |
| 20528 | 23112 | 0.36 | 0.0825 | 1.28E-05 | 1.20E-04 | female |
| 20488 | 23105 | 0.1627 | 0.0373 | 1.30E-05 | 0.000121 | female |
| 20489 | 23112 | -0.135 | 0.0311 | 1.42E-05 | 0.00013 | female |
| 20489 | 23116 | -0.1337 | 0.0308 | 1.44E-05 | 0.000131 | female |
| 20523 | 23100 | 0.192 | 0.0443 | 1.46E-05 | 1.31E-04 | female |
| 20525 | 23124 | -0.2535 | 0.0587 | 1.58E-05 | 1.39E-04 | female |
| 20490 | 23110 | -0.2628 | 0.0609 | 1.58E-05 | 0.000139 | female |
| 20489 | 23120 | -0.1335 | 0.031 | 1.63E-05 | 0.000141 | female |
| 20488 | 23108 | -0.1627 | 0.0378 | 1.70E-05 | 0.000145 | female |
| 20489 | 21001 | -0.1313 | 0.0305 | 1.71E-05 | 0.000145 | female |
| 20528 | 23127 | 0.3326 | 0.0778 | 1.90E-05 | 1.60E-04 | female |
| 20522 | 23111 | -0.2344 | 0.0549 | 1.94E-05 | 1.61E-04 | female |
| 20490 | 23125 | 0.2289 | 0.0537 | 2.03E-05 | 0.000167 | female |
| 20497 | 23111 | 0.195 | 0.0458 | 2.06E-05 | 1.68E-04 | female |
| 20528 | 23099 | 0.3375 | 0.0794 | 2.11E-05 | 1.70E-04 | female |
| 20497 | 21001 | 0.1904 | 0.0448 | 2.14E-05 | 1.71E-04 | female |
| 20521 | 23106 | -0.1606 | 0.0379 | 2.23E-05 | 1.76E-04 | female |
| 20491 | 23111 | -0.2255 | 0.0532 | 2.25E-05 | 0.000176 | female |
| 20497 | 23115 | 0.1967 | 0.0464 | 2.28E-05 | 1.76E-04 | female |
| 20490 | 23106 | -0.2489 | 0.0588 | 2.29E-05 | 0.000176 | female |
| 20488 | 23107 | -0.1645 | 0.0389 | 2.31E-05 | 0.000176 | female |
| 20525 | 23100 | -0.2381 | 0.0564 | 2.42E-05 | 1.82E-04 | female |
| 20489 | 23124 | -0.129 | 0.0306 | 2.45E-05 | 0.000183 | female |
| 20521 | 23119 | 0.1579 | 0.0374 | 2.48E-05 | 1.83E-04 | female |
| 20528 | 23125 | 0.3259 | 0.0776 | 2.67E-05 | 1.95E-04 | female |
| 20490 | 49 | 0.2309 | 0.0552 | 2.90E-05 | 0.00021 | female |
| 20497 | 23104 | 0.1865 | 0.0447 | 3.01E-05 | 2.16E-04 | female |
| 20523 | 23110 | -0.1837 | 0.0441 | 3.09E-05 | 2.20E-04 | female |
| PTSD | 23111 | 0.2245 | 0.0552 | 4.74E-05 | 2.26E-04 | female |
| 20528 | 49 | 0.3358 | 0.0809 | 3.28E-05 | 2.30E-04 | female |
| 20521 | 23116 | 0.1633 | 0.0393 | 3.29E-05 | 2.30E-04 | female |
| 20522 | 48 | -0.2276 | 0.0548 | 3.31E-05 | 2.30E-04 | female |
| 20490 | 23109 | -0.2557 | 0.0616 | 3.35E-05 | 0.00023 | female |
| 20490 | 23099 | 0.255 | 0.0615 | 3.38E-05 | 0.000231 | female |
| 20528 | 23126 | 0.3136 | 0.0757 | 3.46E-05 | 2.34E-04 | female |
| 20528 | 21001 | 0.3435 | 0.0831 | 3.57E-05 | 2.39E-04 | female |
| 20521 | 23112 | 0.1628 | 0.0394 | 3.60E-05 | 2.39E-04 | female |
| 20528 | 23119 | 0.3335 | 0.0809 | 3.73E-05 | 2.45E-04 | female |
| 20489 | 23111 | -0.1282 | 0.0311 | 3.75E-05 | 0.000245 | female |
| 20528 | 23105 | 0.3158 | 0.0766 | 3.78E-05 | 2.45E-04 | female |
| 20491 | 23115 | -0.2181 | 0.0529 | 3.80E-05 | 0.000245 | female |
| 20522 | 23115 | -0.2272 | 0.0552 | 3.92E-05 | 2.48E-04 | female |
| 20490 | 23105 | 0.217 | 0.0528 | 3.95E-05 | 0.000248 | female |
| 20528 | 23104 | 0.3373 | 0.0821 | 3.95E-05 | 2.48E-04 | female |
| 20490 | 23126 | 0.2247 | 0.0547 | 3.97E-05 | 0.000248 | female |
| 20528 | 23123 | 0.333 | 0.0813 | 4.18E-05 | 2.58E-04 | female |
| 20521 | 23120 | 0.1566 | 0.0382 | 4.21E-05 | 2.58E-04 | female |
| 20489 | 48 | -0.1367 | 0.0334 | 4.26E-05 | 0.000258 | female |
| 20490 | 23128 | 0.2283 | 0.0558 | 4.26E-05 | 0.000258 | female |
| 20489 | 23115 | -0.1256 | 0.0308 | 4.64E-05 | 0.000278 | female |
| 20498 | 21002 | 0.1575 | 0.0388 | 4.96E-05 | 2.95E-04 | female |
| 20528 | 23114 | 0.3084 | 0.0761 | 5.11E-05 | 3.02E-04 | female |
| 20528 | 23113 | 0.3076 | 0.0761 | 5.28E-05 | 3.09E-04 | female |
| 20488 | 23102 | 0.1495 | 0.037 | 5.31E-05 | 0.000309 | female |
| 20497 | 23119 | 0.1843 | 0.0457 | 5.45E-05 | 3.15E-04 | female |
| 20528 | 23118 | 0.3071 | 0.0763 | 5.75E-05 | 3.30E-04 | female |
| 20489 | 23119 | -0.1238 | 0.0308 | 5.81E-05 | 0.000331 | female |
| 20528 | 23117 | 0.3075 | 0.0766 | 5.98E-05 | 3.38E-04 | female |
| 20498 | 23110 | -0.1588 | 0.0396 | 6.14E-05 | 3.45E-04 | female |
| 20522 | 23099 | -0.2137 | 0.0534 | 6.32E-05 | 3.53E-04 | female |
| 20498 | 23098 | 0.1557 | 0.039 | 6.49E-05 | 3.60E-04 | female |
| 20525 | 48 | -0.2371 | 0.0594 | 6.60E-05 | 3.63E-04 | female |
| 20521 | 23124 | 0.1529 | 0.0384 | 6.66E-05 | 3.64E-04 | female |
| 20488 | 23128 | 0.1521 | 0.0382 | 6.74E-05 | 0.000366 | female |
| 20497 | 23123 | 0.1821 | 0.0458 | 6.97E-05 | 3.76E-04 | female |
| 20497 | 23112 | 0.1814 | 0.0458 | 7.41E-05 | 3.97E-04 | female |
| PTSD | 23104 | 0.2121 | 0.0549 | 0.0001 | 0.00041 | female |
| PTSD | 23115 | 0.2125 | 0.0547 | 0.0001 | 0.00041 | female |
| 20488 | 23121 | 0.154 | 0.039 | 7.83E-05 | 0.000417 | female |
| 20521 | 23123 | 0.1477 | 0.0374 | 7.93E-05 | 4.19E-04 | female |
| 20488 | 23122 | 0.1524 | 0.0386 | 8.02E-05 | 0.000421 | female |
| 20488 | 23101 | 0.1456 | 0.037 | 8.26E-05 | 0.000431 | female |
| 20528 | 23115 | 0.3117 | 0.0792 | 8.31E-05 | 4.31E-04 | female |
| 20521 | 23109 | -0.1585 | 0.0405 | 9.05E-05 | 4.59E-04 | female |
| 20498 | 23106 | -0.1527 | 0.039 | 9.11E-05 | 4.59E-04 | female |
| 20489 | 23123 | -0.1186 | 0.0303 | 9.20E-05 | 0.000459 | female |
| 20497 | 23116 | 0.1806 | 0.0464 | 9.83E-05 | 4.59E-04 | female |
| 20489 | 23098 | -0.1183 | 0.0304 | 9.90E-05 | 0.000459 | female |
| 20488 | 23117 | 0.1406 | 0.0362 | 0.0001 | 0.000459 | female |
| 20488 | 23118 | 0.14 | 0.0361 | 0.0001 | 0.000459 | female |
| 20488 | 23130 | 0.1453 | 0.0379 | 0.0001 | 0.000459 | female |
| 20490 | 23117 | 0.1976 | 0.0518 | 0.0001 | 0.000459 | female |
| 20490 | 23118 | 0.1987 | 0.0519 | 0.0001 | 0.000459 | female |
| 20495 | 23119 | 0.1839 | 0.0483 | 0.0001 | 0.000459 | female |
| 20497 | 23120 | 0.1773 | 0.0457 | 0.0001 | 0.000459 | female |
| 20497 | 23124 | 0.176 | 0.0454 | 0.0001 | 0.000459 | female |
| 20522 | 23127 | -0.1995 | 0.052 | 0.0001 | 0.000459 | female |
| 20523 | 23106 | -0.1689 | 0.0439 | 0.0001 | 0.000459 | female |
| 20525 | 23127 | -0.2134 | 0.0555 | 0.0001 | 0.000459 | female |
| 20525 | 23128 | -0.2131 | 0.0549 | 0.0001 | 0.000459 | female |
| 20528 | 23101 | 0.2843 | 0.0744 | 0.0001 | 0.000459 | female |
| 20528 | 23102 | 0.2829 | 0.0741 | 0.0001 | 0.000459 | female |
| 20528 | 23111 | 0.3035 | 0.079 | 0.0001 | 0.000459 | female |
| PTSD | 23119 | 0.2071 | 0.056 | 0.0002 | 0.000782 | female |
| 20487 | 21001 | 0.1622 | 0.0434 | 0.0002 | 0.000814 | female |
| 20488 | 23114 | 0.1345 | 0.0365 | 0.0002 | 0.000814 | female |
| 20488 | 23129 | 0.1431 | 0.0378 | 0.0002 | 0.000814 | female |
| 20489 | 23100 | -0.1154 | 0.0306 | 0.0002 | 0.000814 | female |
| 20489 | 23110 | 0.1181 | 0.0319 | 0.0002 | 0.000814 | female |
| 20490 | 23108 | -0.2132 | 0.058 | 0.0002 | 0.000814 | female |
| 20490 | 23113 | 0.1937 | 0.0526 | 0.0002 | 0.000814 | female |
| 20490 | 23114 | 0.1938 | 0.0526 | 0.0002 | 0.000814 | female |
| 20490 | 23121 | 0.2046 | 0.0548 | 0.0002 | 0.000814 | female |
| 20490 | 23122 | 0.2003 | 0.0541 | 0.0002 | 0.000814 | female |
| 20498 | 23128 | 0.1469 | 0.0397 | 0.0002 | 0.000814 | female |
| 20521 | 48 | 0.157 | 0.0424 | 0.0002 | 0.000814 | female |
| 20522 | 23100 | -0.1934 | 0.0526 | 0.0002 | 0.000814 | female |
| 20523 | 23109 | -0.1653 | 0.0437 | 0.0002 | 0.000814 | female |
| 20523 | 23127 | 0.1658 | 0.0439 | 0.0002 | 0.000814 | female |
| 20523 | 23128 | 0.1636 | 0.0443 | 0.0002 | 0.000814 | female |
| 20525 | 21002 | -0.21 | 0.0557 | 0.0002 | 0.000814 | female |
| 20525 | 23098 | -0.2067 | 0.0563 | 0.0002 | 0.000814 | female |
| 20525 | 49 | -0.2034 | 0.0545 | 0.0002 | 0.000814 | female |
| 20528 | 23121 | 0.2802 | 0.0752 | 0.0002 | 0.000814 | female |
| 20528 | 23122 | 0.2788 | 0.0746 | 0.0002 | 0.000814 | female |
| 20489 | 21002 | -0.1146 | 0.0307 | 0.0002 | 0.000814 | female |
| PTSD | 23112 | 0.1899 | 0.0524 | 0.0003 | 0.001075 | female |
| PTSD | 23116 | 0.1879 | 0.0522 | 0.0003 | 0.001075 | female |
| 20487 | 23104 | 0.1579 | 0.0435 | 0.0003 | 0.001103 | female |
| 20487 | 23110 | -0.1426 | 0.0396 | 0.0003 | 0.001103 | female |
| 20488 | 23113 | 0.1341 | 0.0367 | 0.0003 | 0.001103 | female |
| 20489 | 23125 | -0.1137 | 0.0315 | 0.0003 | 0.001103 | female |
| 20490 | 23101 | 0.191 | 0.0527 | 0.0003 | 0.001103 | female |
| 20490 | 23102 | 0.1932 | 0.0528 | 0.0003 | 0.001103 | female |
| 20490 | 23107 | -0.216 | 0.0591 | 0.0003 | 0.001103 | female |
| 20495 | 21001 | 0.1779 | 0.049 | 0.0003 | 0.001103 | female |
| 20495 | 23104 | 0.1748 | 0.0486 | 0.0003 | 0.001103 | female |
| 20495 | 23120 | 0.1783 | 0.0491 | 0.0003 | 0.001103 | female |
| 20497 | 23099 | 0.1653 | 0.0459 | 0.0003 | 0.001103 | female |
| 20497 | 48 | 0.1678 | 0.0468 | 0.0003 | 0.001103 | female |
| 20498 | 23109 | -0.1407 | 0.0392 | 0.0003 | 0.001103 | female |
| 20498 | 23127 | 0.1461 | 0.0407 | 0.0003 | 0.001103 | female |
| 20498 | 50 | -0.1107 | 0.0307 | 0.0003 | 0.001103 | female |
| 20522 | 23112 | -0.1961 | 0.0538 | 0.0003 | 0.001103 | female |
| 20522 | 23116 | -0.194 | 0.0539 | 0.0003 | 0.001103 | female |
| 20522 | 23119 | -0.1926 | 0.0527 | 0.0003 | 0.001103 | female |
| 20522 | 23128 | -0.1855 | 0.0513 | 0.0003 | 0.001103 | female |
| 20523 | 21002 | 0.165 | 0.0455 | 0.0003 | 0.001103 | female |
| 20523 | 23098 | 0.1663 | 0.0456 | 0.0003 | 0.001103 | female |
| PTSD | 23123 | 0.1968 | 0.0559 | 0.0004 | 0.001323 | female |
| 20489 | 23106 | 0.1096 | 0.031 | 0.0004 | 0.001412 | female |
| 20490 | 23127 | 0.2046 | 0.0581 | 0.0004 | 0.001412 | female |
| 20491 | 21001 | -0.1973 | 0.0562 | 0.0004 | 0.001412 | female |
| 20491 | 23104 | -0.2004 | 0.0565 | 0.0004 | 0.001412 | female |
| 20495 | 23123 | 0.1698 | 0.0476 | 0.0004 | 0.001412 | female |
| 20495 | 23124 | 0.1727 | 0.0486 | 0.0004 | 0.001412 | female |
| 20497 | 23100 | 0.1621 | 0.0459 | 0.0004 | 0.001412 | female |
| 20522 | 23123 | -0.1872 | 0.0527 | 0.0004 | 0.001412 | female |
| 20528 | 23130 | 0.2568 | 0.0732 | 0.0004 | 0.001412 | female |
| PTSD | 23120 | 0.1847 | 0.0534 | 0.0005 | 0.001536 | female |
| PTSD | 23124 | 0.1862 | 0.0533 | 0.0005 | 0.001536 | female |
| 20487 | 23111 | 0.1525 | 0.0436 | 0.0005 | 0.00172 | female |
| 20489 | 23099 | -0.1069 | 0.0309 | 0.0005 | 0.00172 | female |
| 20489 | 23105 | -0.1076 | 0.0307 | 0.0005 | 0.00172 | female |
| 20498 | 49 | 0.136 | 0.0392 | 0.0005 | 0.00172 | female |
| 20522 | 23120 | -0.1853 | 0.0531 | 0.0005 | 0.00172 | female |
| 20528 | 23129 | 0.256 | 0.0731 | 0.0005 | 0.00172 | female |
| PTSD | 23099 | 0.1874 | 0.0548 | 0.0006 | 0.00172 | female |
| 20489 | 23126 | -0.1095 | 0.0318 | 0.0006 | 0.00202 | female |
| 20491 | 23112 | -0.1906 | 0.0555 | 0.0006 | 0.00202 | female |
| 20495 | 23112 | 0.1677 | 0.0488 | 0.0006 | 0.00202 | female |
| 20522 | 23124 | -0.1804 | 0.0529 | 0.0006 | 0.00202 | female |
| 20530 | 21002 | 0.256 | 0.0743 | 0.0006 | 0.00202 | female |
| 20487 | 23106 | -0.1306 | 0.0387 | 0.0007 | 0.002308 | female |
| 20487 | 23115 | 0.1483 | 0.0438 | 0.0007 | 0.002308 | female |
| 20488 | 23127 | 0.1303 | 0.0383 | 0.0007 | 0.002308 | female |
| 20491 | 23116 | -0.1876 | 0.0555 | 0.0007 | 0.002308 | female |
| 20522 | 21001 | -0.1787 | 0.0528 | 0.0007 | 0.002308 | female |
| 20495 | 23111 | 0.1616 | 0.0482 | 0.0008 | 0.002595 | female |
| 20495 | 23116 | 0.165 | 0.0492 | 0.0008 | 0.002595 | female |
| 20530 | 23098 | 0.2499 | 0.0745 | 0.0008 | 0.002595 | female |
| 20530 | 23120 | 0.2636 | 0.0786 | 0.0008 | 0.002595 | female |
| 20489 | 23128 | -0.1008 | 0.0303 | 0.0009 | 0.002884 | female |
| 20491 | 23119 | -0.1797 | 0.0543 | 0.0009 | 0.002884 | female |
| 20498 | 23107 | -0.1277 | 0.0385 | 0.0009 | 0.002884 | female |
| 20487 | 48 | 0.1491 | 0.0454 | 0.001 | 0.003117 | female |
| 20490 | 23129 | 0.1762 | 0.0533 | 0.001 | 0.003117 | female |
| 20490 | 23130 | 0.1769 | 0.0536 | 0.001 | 0.003117 | female |
| 20491 | 23123 | -0.1792 | 0.0542 | 0.001 | 0.003117 | female |
| 20522 | 23104 | -0.1718 | 0.0524 | 0.001 | 0.003117 | female |
| 20530 | 23100 | 0.2441 | 0.0744 | 0.001 | 0.003117 | female |
| 20530 | 23116 | 0.2499 | 0.0761 | 0.001 | 0.003117 | female |
| PTSD | 23100 | 0.1678 | 0.0517 | 0.0012 | 0.003225 | female |
| 20487 | 23112 | 0.1428 | 0.0438 | 0.0011 | 0.003389 | female |
| 20489 | 23117 | -0.0985 | 0.0302 | 0.0011 | 0.003389 | female |
| 20530 | 21001 | 0.2558 | 0.0782 | 0.0011 | 0.003389 | female |
| 20487 | 23116 | 0.142 | 0.044 | 0.0012 | 0.003627 | female |
| 20487 | 23119 | 0.1374 | 0.0425 | 0.0012 | 0.003627 | female |
| 20495 | 23115 | 0.1572 | 0.0486 | 0.0012 | 0.003627 | female |
| 20530 | 23124 | 0.2495 | 0.0769 | 0.0012 | 0.003627 | female |
| 20530 | 49 | 0.2518 | 0.078 | 0.0012 | 0.003627 | female |
| 20487 | 23120 | 0.1381 | 0.0429 | 0.0013 | 0.003857 | female |
| 20489 | 23118 | -0.0969 | 0.0302 | 0.0013 | 0.003857 | female |
| 20491 | 23120 | -0.1793 | 0.0556 | 0.0013 | 0.003857 | female |
| 20521 | 23100 | 0.1257 | 0.039 | 0.0013 | 0.003857 | female |
| 20530 | 23112 | 0.2452 | 0.0763 | 0.0013 | 0.003857 | female |
| 20497 | 21002 | 0.145 | 0.0454 | 0.0014 | 0.004107 | female |
| 20530 | 23104 | 0.2491 | 0.078 | 0.0014 | 0.004107 | female |
| 20530 | 23119 | 0.2467 | 0.0772 | 0.0014 | 0.004107 | female |
| 20489 | 23101 | -0.0984 | 0.0309 | 0.0015 | 0.004337 | female |
| 20489 | 23102 | -0.0983 | 0.0309 | 0.0015 | 0.004337 | female |
| 20491 | 23124 | -0.1769 | 0.0556 | 0.0015 | 0.004337 | female |
| 20523 | 23107 | -0.1453 | 0.0459 | 0.0015 | 0.004337 | female |
| 20487 | 23124 | 0.136 | 0.043 | 0.0016 | 0.004544 | female |
| 20495 | 23100 | 0.1518 | 0.0482 | 0.0016 | 0.004544 | female |
| 20521 | 23108 | -0.1171 | 0.0372 | 0.0016 | 0.004544 | female |
| 20523 | 49 | 0.1383 | 0.0437 | 0.0016 | 0.004544 | female |
| 20523 | 50 | -0.1271 | 0.0403 | 0.0016 | 0.004544 | female |
| 20489 | 23109 | 0.1002 | 0.0319 | 0.0017 | 0.004727 | female |
| 20489 | 23121 | -0.0978 | 0.0312 | 0.0017 | 0.004727 | female |
| 20497 | 23098 | 0.1424 | 0.0453 | 0.0017 | 0.004727 | female |
| 20498 | 23125 | 0.1168 | 0.0371 | 0.0017 | 0.004727 | female |
| 20521 | 23099 | 0.1211 | 0.0386 | 0.0017 | 0.004727 | female |
| 20530 | 23126 | 0.2303 | 0.0734 | 0.0017 | 0.004727 | female |
| 20496 | 23110 | -0.2218 | 0.0711 | 0.0018 | 0.00497 | female |
| 20498 | 23108 | -0.1223 | 0.0392 | 0.0018 | 0.00497 | female |
| 20489 | 23122 | -0.0975 | 0.0315 | 0.0019 | 0.005228 | female |
| 20487 | 23123 | 0.1323 | 0.0428 | 0.002 | 0.005447 | female |
| 20491 | 23099 | -0.1629 | 0.0527 | 0.002 | 0.005447 | female |
| 20530 | 23105 | 0.2268 | 0.0735 | 0.002 | 0.005447 | female |
| 20489 | 49 | -0.1014 | 0.0329 | 0.0021 | 0.005605 | female |
| 20497 | 23128 | 0.1392 | 0.0452 | 0.0021 | 0.005605 | female |
| 20522 | 21002 | -0.1555 | 0.0505 | 0.0021 | 0.005605 | female |
| 20522 | 49 | -0.1615 | 0.0525 | 0.0021 | 0.005605 | female |
| 20523 | 23108 | -0.1435 | 0.0467 | 0.0021 | 0.005605 | female |
| 20530 | 23128 | 0.2204 | 0.0717 | 0.0021 | 0.005605 | female |
| 20489 | 23113 | -0.0924 | 0.0302 | 0.0022 | 0.005813 | female |
| 20489 | 23114 | -0.0927 | 0.0302 | 0.0022 | 0.005813 | female |
| 20530 | 23125 | 0.2241 | 0.0731 | 0.0022 | 0.005813 | female |
| 20487 | 23109 | -0.1223 | 0.0401 | 0.0023 | 0.006018 | female |
| 20489 | 23130 | -0.097 | 0.0318 | 0.0023 | 0.006018 | female |
| 20530 | 23123 | 0.2306 | 0.0757 | 0.0023 | 0.006018 | female |
| 20521 | 23098 | 0.1193 | 0.0393 | 0.0024 | 0.006238 | female |
| 20525 | 23110 | 0.1746 | 0.0575 | 0.0024 | 0.006238 | female |
| 20495 | 49 | 0.1463 | 0.0483 | 0.0025 | 0.006456 | female |
| 20521 | 23107 | -0.1116 | 0.0369 | 0.0025 | 0.006456 | female |
| 20489 | 23129 | -0.0953 | 0.0318 | 0.0027 | 0.006906 | female |
| 20495 | 21002 | 0.1453 | 0.0484 | 0.0027 | 0.006906 | female |
| 20521 | 21002 | 0.1182 | 0.0394 | 0.0027 | 0.006906 | female |
| 20491 | 48 | -0.1608 | 0.0539 | 0.0028 | 0.007116 | female |
| 20498 | 23105 | 0.1107 | 0.037 | 0.0028 | 0.007116 | female |
| 20495 | 23098 | 0.1425 | 0.0478 | 0.0029 | 0.007347 | female |
| 20528 | 23107 | -0.2244 | 0.0756 | 0.003 | 0.007552 | female |
| 20530 | 23114 | 0.222 | 0.0747 | 0.003 | 0.007552 | female |
| 20522 | 23098 | -0.1491 | 0.0504 | 0.0031 | 0.007755 | female |
| 20531 | 23107 | -0.1438 | 0.0487 | 0.0031 | 0.007755 | female |
| 20498 | 23126 | 0.1089 | 0.037 | 0.0032 | 0.007906 | female |
| 20530 | 23113 | 0.2199 | 0.0747 | 0.0032 | 0.007906 | female |
| 20530 | 23115 | 0.2164 | 0.0735 | 0.0032 | 0.007906 | female |
| 20531 | 23106 | -0.1363 | 0.0463 | 0.0032 | 0.007906 | female |
| 20491 | 23110 | 0.1703 | 0.058 | 0.0033 | 0.008078 | female |
| 20495 | 23099 | 0.1389 | 0.0473 | 0.0033 | 0.008078 | female |
| 20495 | 48 | 0.1556 | 0.053 | 0.0033 | 0.008078 | female |
| 20530 | 23099 | 0.2134 | 0.0729 | 0.0034 | 0.008297 | female |
| PTSD | 23128 | 0.1521 | 0.0518 | 0.0034 | 0.0086 | female |
| 20497 | 23127 | 0.1328 | 0.0457 | 0.0036 | 0.008759 | female |
| PTSD | 49 | 0.1525 | 0.0524 | 0.0036 | 0.008846 | female |
| 20489 | 23108 | 0.0884 | 0.0306 | 0.0039 | 0.009374 | female |
| 20530 | 23102 | 0.2108 | 0.0729 | 0.0039 | 0.009374 | female |
| 20530 | 23118 | 0.2144 | 0.0743 | 0.0039 | 0.009374 | female |
| 20531 | 23108 | -0.1393 | 0.0483 | 0.0039 | 0.009374 | female |
| 20530 | 23101 | 0.2088 | 0.0727 | 0.0041 | 0.009796 | female |
| 20531 | 21001 | 0.1337 | 0.0466 | 0.0041 | 0.009796 | female |
| 20497 | 23110 | -0.126 | 0.044 | 0.0042 | 0.010005 | female |
| PTSD | 23127 | 0.1559 | 0.0545 | 0.0042 | 0.010033 | female |
| 20496 | 23106 | -0.2003 | 0.0702 | 0.0043 | 0.010182 | female |
| 20530 | 23117 | 0.2124 | 0.0745 | 0.0043 | 0.010182 | female |
| 20525 | 23125 | -0.1583 | 0.0559 | 0.0046 | 0.01086 | female |
| 20491 | 23100 | -0.151 | 0.0534 | 0.0047 | 0.011064 | female |
| 20489 | 23127 | -0.087 | 0.0309 | 0.0048 | 0.011233 | female |
| 20491 | 23109 | 0.1628 | 0.0577 | 0.0048 | 0.011233 | female |
| 20521 | 23126 | 0.1142 | 0.0407 | 0.005 | 0.011633 | female |
| 20530 | 23111 | 0.2081 | 0.0741 | 0.005 | 0.011633 | female |
| 20495 | 23106 | -0.1352 | 0.0486 | 0.0054 | 0.01249 | female |
| 20530 | 23130 | 0.2027 | 0.0728 | 0.0054 | 0.01249 | female |
| PTSD | 23098 | 0.1371 | 0.0499 | 0.006 | 0.012585 | female |
| 20489 | 23107 | 0.0857 | 0.0309 | 0.0055 | 0.012685 | female |
| 20491 | 50 | 0.1167 | 0.0422 | 0.0057 | 0.013108 | female |
| 20521 | 23125 | 0.1108 | 0.0402 | 0.0058 | 0.013262 | female |
| 20531 | 21002 | 0.1269 | 0.046 | 0.0058 | 0.013262 | female |
| 20521 | 49 | 0.1094 | 0.0398 | 0.0059 | 0.013452 | female |
| 20530 | 23129 | 0.1983 | 0.0722 | 0.006 | 0.013641 | female |
| 20528 | 23108 | -0.2105 | 0.0767 | 0.0061 | 0.013829 | female |
| 20531 | 23104 | 0.1269 | 0.0464 | 0.0063 | 0.014242 | female |
| 20525 | 23126 | -0.1528 | 0.056 | 0.0064 | 0.014427 | female |
| 20495 | 23108 | -0.1327 | 0.049 | 0.0068 | 0.0152 | female |
| 20528 | 23106 | -0.1996 | 0.0738 | 0.0068 | 0.0152 | female |
| 20530 | 23127 | 0.1916 | 0.0708 | 0.0068 | 0.0152 | female |
| 20495 | 23125 | 0.1274 | 0.0472 | 0.0069 | 0.01538 | female |
| 20531 | 23120 | 0.1262 | 0.0469 | 0.0071 | 0.015782 | female |
| 20496 | 23109 | -0.194 | 0.0723 | 0.0073 | 0.016182 | female |
| 20531 | 23114 | 0.1258 | 0.0471 | 0.0075 | 0.016579 | female |
| 20531 | 23117 | 0.1241 | 0.0466 | 0.0077 | 0.016974 | female |
| 20531 | 23118 | 0.1239 | 0.0466 | 0.0079 | 0.017367 | female |
| 20487 | 23100 | 0.1123 | 0.0424 | 0.0081 | 0.017709 | female |
| 20531 | 23113 | 0.1241 | 0.0469 | 0.0081 | 0.017709 | female |
| 20495 | 23105 | 0.125 | 0.0473 | 0.0083 | 0.018097 | female |
| 20495 | 23128 | 0.1232 | 0.0469 | 0.0087 | 0.018917 | female |
| 20530 | 23122 | 0.1837 | 0.0701 | 0.0088 | 0.019083 | female |
| 20495 | 23107 | -0.1272 | 0.0486 | 0.0089 | 0.019247 | female |
| 20525 | 23109 | 0.152 | 0.0582 | 0.0091 | 0.019574 | female |
| 20531 | 23098 | 0.1196 | 0.0458 | 0.0091 | 0.019574 | female |
| 20521 | 23105 | 0.1007 | 0.0389 | 0.0095 | 0.02027 | female |
| 20521 | 50 | -0.0942 | 0.0363 | 0.0095 | 0.02027 | female |
| 20530 | 23121 | 0.1839 | 0.071 | 0.0095 | 0.02027 | female |
| 20497 | 23109 | -0.1116 | 0.0431 | 0.0096 | 0.020374 | female |
| 20531 | 23105 | 0.1206 | 0.0466 | 0.0096 | 0.020374 | female |
| 20487 | 21002 | 0.1068 | 0.0413 | 0.0097 | 0.020532 | female |
| 20498 | 23118 | 0.0949 | 0.0368 | 0.0098 | 0.020689 | female |
| 20531 | 23116 | 0.1196 | 0.0464 | 0.0099 | 0.020845 | female |
| 20487 | 23099 | 0.1093 | 0.0425 | 0.0101 | 0.021099 | female |
| 20524 | 23110 | -0.1451 | 0.0564 | 0.0101 | 0.021099 | female |
| 20525 | 23105 | -0.1421 | 0.0552 | 0.0101 | 0.021099 | female |
| 20487 | 23108 | -0.1012 | 0.0395 | 0.0103 | 0.021405 | female |
| 20497 | 23106 | -0.1115 | 0.0434 | 0.0103 | 0.021405 | female |
| 20531 | 23112 | 0.1182 | 0.0461 | 0.0104 | 0.021556 | female |
| 20523 | 23126 | 0.1191 | 0.0466 | 0.0106 | 0.021914 | female |
| 20496 | 23126 | 0.1602 | 0.0629 | 0.0108 | 0.022212 | female |
| 20497 | 49 | 0.1218 | 0.0478 | 0.0108 | 0.022212 | female |
| 20528 | 23110 | -0.1866 | 0.0733 | 0.011 | 0.022566 | female |
| 20487 | 49 | 0.1071 | 0.0422 | 0.0111 | 0.022712 | female |
| 20495 | 23117 | 0.1195 | 0.0471 | 0.0112 | 0.022742 | female |
| 20498 | 23117 | 0.0932 | 0.0367 | 0.0112 | 0.022742 | female |
| 20531 | 23110 | -0.1211 | 0.0477 | 0.0112 | 0.022742 | female |
| 20487 | 23098 | 0.1046 | 0.0414 | 0.0115 | 0.023233 | female |
| 20531 | 49 | 0.1115 | 0.0442 | 0.0115 | 0.023233 | female |
| 20523 | 23105 | 0.1155 | 0.046 | 0.012 | 0.024182 | female |
| PTSD | 23110 | -0.1404 | 0.0563 | 0.0127 | 0.024271 | female |
| 20495 | 23118 | 0.118 | 0.047 | 0.0121 | 0.024322 | female |
| 20495 | 23110 | -0.1228 | 0.0491 | 0.0124 | 0.024862 | female |
| 20523 | 23125 | 0.1152 | 0.0461 | 0.0125 | 0.025 | female |
| 20496 | 23125 | 0.1557 | 0.0624 | 0.0126 | 0.025137 | female |
| 20525 | 23106 | 0.1416 | 0.057 | 0.0129 | 0.025671 | female |
| 20531 | 23102 | 0.116 | 0.0468 | 0.0132 | 0.026203 | female |
| 20498 | 23102 | 0.0899 | 0.0363 | 0.0133 | 0.026336 | female |
| 20531 | 48 | 0.1224 | 0.0496 | 0.0136 | 0.026863 | female |
| 20531 | 23101 | 0.1154 | 0.047 | 0.014 | 0.027585 | female |
| 20524 | 50 | -0.1159 | 0.0474 | 0.0144 | 0.028234 | female |
| 20530 | 23107 | -0.1982 | 0.081 | 0.0144 | 0.028234 | female |
| 20495 | 23126 | 0.1153 | 0.0474 | 0.0149 | 0.029071 | female |
| 20531 | 23124 | 0.1131 | 0.0464 | 0.0149 | 0.029071 | female |
| 20530 | 23106 | -0.1917 | 0.0788 | 0.015 | 0.029195 | female |
| 20495 | 23102 | 0.1142 | 0.047 | 0.0152 | 0.029512 | female |
| 20498 | 23114 | 0.0876 | 0.0362 | 0.0154 | 0.029828 | female |
| 20524 | 23106 | -0.1357 | 0.0561 | 0.0156 | 0.03007 | female |
| 20531 | 23119 | 0.1152 | 0.0476 | 0.0156 | 0.03007 | female |
| PTSD | BMI | 0.1344 | 0.0559 | 0.0161 | 0.0301 | female |
| 20497 | 23126 | 0.1112 | 0.046 | 0.0157 | 0.030189 | female |
| 20496 | 21001 | 0.1518 | 0.0629 | 0.0158 | 0.030309 | female |
| 20498 | 23113 | 0.0874 | 0.0363 | 0.0159 | 0.030427 | female |
| 20497 | 23125 | 0.1099 | 0.0457 | 0.0161 | 0.030736 | female |
| 20531 | 23126 | 0.1143 | 0.0475 | 0.0162 | 0.030853 | female |
| 20491 | 23098 | -0.1279 | 0.0533 | 0.0165 | 0.03135 | female |
| 20495 | 23114 | 0.1125 | 0.047 | 0.0166 | 0.031465 | female |
| 20496 | 23104 | 0.1504 | 0.0628 | 0.0167 | 0.03158 | female |
| 20528 | 23109 | -0.175 | 0.0732 | 0.0169 | 0.031882 | female |
| 20498 | 23101 | 0.0868 | 0.0364 | 0.017 | 0.031995 | female |
| 20487 | 23125 | 0.0945 | 0.0396 | 0.0171 | 0.032108 | female |
| 20495 | 23113 | 0.1124 | 0.0472 | 0.0172 | 0.03222 | female |
| 20531 | 23100 | 0.1098 | 0.0462 | 0.0174 | 0.032518 | female |
| 20496 | 23102 | 0.1411 | 0.0595 | 0.0177 | 0.033001 | female |
| 20495 | 23101 | 0.1117 | 0.0471 | 0.0178 | 0.033033 | female |
| 20524 | 23109 | -0.1292 | 0.0545 | 0.0178 | 0.033033 | female |
| 20496 | 23130 | 0.1461 | 0.0617 | 0.0179 | 0.033142 | female |
| 20487 | 23107 | -0.0957 | 0.0405 | 0.0181 | 0.033435 | female |
| 20497 | 23105 | 0.1066 | 0.0451 | 0.0182 | 0.033542 | female |
| 20531 | 23125 | 0.1102 | 0.0467 | 0.0184 | 0.033832 | female |
| 20487 | 50 | -0.0774 | 0.0329 | 0.0185 | 0.033938 | female |
| 20530 | 48 | 0.1754 | 0.0747 | 0.0188 | 0.034409 | female |
| 20491 | 23128 | -0.1222 | 0.052 | 0.0189 | 0.034434 | female |
| 20521 | 23128 | 0.0924 | 0.0394 | 0.0189 | 0.034434 | female |
| 20531 | 23123 | 0.1108 | 0.0474 | 0.0194 | 0.035265 | female |
| 20498 | 23121 | 0.0851 | 0.0365 | 0.0196 | 0.035547 | female |
| 20487 | 23126 | 0.093 | 0.0399 | 0.02 | 0.03619 | female |
| 20495 | 23127 | 0.1087 | 0.0468 | 0.0203 | 0.036567 | female |
| 20521 | 23102 | 0.0896 | 0.0386 | 0.0203 | 0.036567 | female |
| 20496 | 23101 | 0.139 | 0.06 | 0.0205 | 0.036762 | female |
| 20496 | 23129 | 0.1428 | 0.0616 | 0.0205 | 0.036762 | female |
| 20488 | 50 | -0.0801 | 0.0347 | 0.021 | 0.037574 | female |
| PTSD | 23109 | -0.1285 | 0.0556 | 0.0208 | 0.03806 | female |
| 20529 | 23130 | 0.1517 | 0.0661 | 0.0217 | 0.038653 | female |
| 20530 | 23108 | -0.1871 | 0.0815 | 0.0217 | 0.038653 | female |
| 20491 | 21002 | -0.1215 | 0.053 | 0.0219 | 0.038836 | female |
| 20529 | 23129 | 0.1506 | 0.0657 | 0.0219 | 0.038836 | female |
| 20491 | 23127 | -0.1192 | 0.0521 | 0.022 | 0.038841 | female |
| 20521 | 23130 | 0.0901 | 0.0393 | 0.022 | 0.038841 | female |
| 20496 | 23105 | 0.1371 | 0.0601 | 0.0224 | 0.03946 | female |
| 20521 | 23101 | 0.0881 | 0.0386 | 0.0226 | 0.039724 | female |
| 20495 | 23130 | 0.1088 | 0.0478 | 0.0229 | 0.040163 | female |
| 20498 | 20015 | -0.0782 | 0.0344 | 0.023 | 0.04025 | female |
| 20496 | 23107 | -0.1582 | 0.0697 | 0.0232 | 0.040423 | female |
| 20521 | 23122 | 0.0913 | 0.0402 | 0.0232 | 0.040423 | female |
| 20496 | 23121 | 0.1413 | 0.0624 | 0.0235 | 0.040767 | female |
| 20521 | 23129 | 0.0892 | 0.0394 | 0.0235 | 0.040767 | female |
| PTSD | 23106 | -0.123 | 0.0544 | 0.0238 | 0.040936 | female |
| 20496 | 23122 | 0.1402 | 0.0621 | 0.0239 | 0.041371 | female |
| 20494 | 48 | 0.1534 | 0.0681 | 0.0242 | 0.0418 | female |
| 20495 | 23129 | 0.1071 | 0.0477 | 0.0246 | 0.042399 | female |
| 20531 | 23129 | 0.1064 | 0.0477 | 0.0257 | 0.0442 | female |
| 20531 | 23130 | 0.1062 | 0.0477 | 0.026 | 0.044619 | female |
| 20491 | 23106 | 0.1314 | 0.0591 | 0.0261 | 0.044695 | female |
| 20498 | 23122 | 0.0809 | 0.0365 | 0.0266 | 0.045454 | female |
| 20531 | 23109 | -0.1055 | 0.0476 | 0.0268 | 0.045697 | female |
| 20487 | 23105 | 0.0851 | 0.0387 | 0.0277 | 0.047131 | female |
| 20523 | 23118 | 0.1027 | 0.0467 | 0.0278 | 0.047201 | female |
| 20525 | 23121 | -0.1244 | 0.0567 | 0.0282 | 0.047778 | female |
| 20523 | 23117 | 0.1015 | 0.0466 | 0.0294 | 0.049706 | female |
| 20527 | 23111 | 0.4051 | 0.0649 | 4.42E-10 | 1.36E-07 | male |
| 20527 | 23115 | 0.4057 | 0.0651 | 4.7E-10 | 1.36E-07 | male |
| 20527 | 21001 | 0.4085 | 0.066 | 6.03E-10 | 1.36E-07 | male |
| 20527 | 23112 | 0.3969 | 0.0646 | 8.21E-10 | 1.36E-07 | male |
| 20527 | 23104 | 0.4125 | 0.0672 | 8.55E-10 | 1.36E-07 | male |
| 20527 | 23116 | 0.3922 | 0.065 | 1.56E-09 | 2.07E-07 | male |
| 20488 | 23111 | 0.2845 | 0.0474 | 1.96E-09 | 2.23E-07 | male |
| 20527 | 23099 | 0.3781 | 0.0645 | 4.52E-09 | 4.49E-07 | male |
| 20488 | 21001 | 0.2877 | 0.0494 | 5.6E-09 | 4.49E-07 | male |
| 20488 | 23104 | 0.2841 | 0.0489 | 6.11E-09 | 4.49E-07 | male |
| 20488 | 23112 | 0.2785 | 0.0479 | 6.19E-09 | 4.49E-07 | male |
| 20488 | 23115 | 0.2745 | 0.0479 | 1.01E-08 | 6.72E-07 | male |
| 20527 | 23100 | 0.3709 | 0.0656 | 1.54E-08 | 9.45E-07 | male |
| 20527 | 23127 | 0.3623 | 0.0645 | 1.93E-08 | 1.1E-06 | male |
| 20488 | 23116 | 0.2662 | 0.0483 | 3.64E-08 | 1.94E-06 | male |
| 20527 | 23128 | 0.359 | 0.0654 | 3.97E-08 | 1.98E-06 | male |
| 20527 | 23123 | 0.3408 | 0.0643 | 1.15E-07 | 5.4E-06 | male |
| 20527 | 48 | 0.3516 | 0.0673 | 1.73E-07 | 7.67E-06 | male |
| 20488 | 23110 | -0.2518 | 0.0484 | 1.94E-07 | 7.75E-06 | male |
| 20527 | 23124 | 0.3431 | 0.066 | 2.03E-07 | 7.75E-06 | male |
| 20530 | 21001 | 0.2938 | 0.0565 | 2.04E-07 | 7.75E-06 | male |
| 20527 | 23119 | 0.3354 | 0.0651 | 2.61E-07 | 9.47E-06 | male |
| 20530 | 23104 | 0.2926 | 0.0569 | 2.75E-07 | 9.54E-06 | male |
| 20527 | 23120 | 0.3381 | 0.066 | 2.99E-07 | 9.94E-06 | male |
| 20488 | 23109 | -0.2416 | 0.0475 | 3.64E-07 | 1.16E-05 | male |
| 20488 | 48 | 0.2476 | 0.0488 | 3.97E-07 | 1.22E-05 | male |
| 20530 | 23116 | 0.2996 | 0.0599 | 5.58E-07 | 1.65E-05 | male |
| 20530 | 23112 | 0.299 | 0.0598 | 5.86E-07 | 1.67E-05 | male |
| 20498 | 23111 | 0.3251 | 0.0658 | 7.65E-07 | 2.11E-05 | male |
| 20530 | 23100 | 0.2865 | 0.0587 | 1.04E-06 | 2.73E-05 | male |
| 20498 | 23115 | 0.3189 | 0.0653 | 1.06E-06 | 2.73E-05 | male |
| 20530 | 23124 | 0.2779 | 0.0572 | 1.18E-06 | 2.94E-05 | male |
| 20488 | 23099 | 0.2259 | 0.0467 | 1.32E-06 | 3.19E-05 | male |
| 20530 | 48 | 0.2853 | 0.0596 | 1.67E-06 | 3.92E-05 | male |
| 20488 | 23100 | 0.2257 | 0.0477 | 2.25E-06 | 5.13E-05 | male |
| 20530 | 21002 | 0.2559 | 0.0546 | 2.74E-06 | 6.07E-05 | male |
| 20530 | 23098 | 0.256 | 0.0547 | 2.88E-06 | 6.21E-05 | male |
| 20498 | 23112 | 0.2996 | 0.0646 | 3.53E-06 | 7.41E-05 | male |
| 20530 | 23120 | 0.2635 | 0.0571 | 3.93E-06 | 8.04E-05 | male |
| 20530 | 23128 | 0.2706 | 0.0594 | 5.13E-06 | 0.000102 | male |
| 20530 | 23115 | 0.2816 | 0.0619 | 5.32E-06 | 0.000104 | male |
| 20498 | 23116 | 0.2921 | 0.0643 | 5.6E-06 | 0.000106 | male |
| 20527 | 23109 | -0.2903 | 0.064 | 5.74E-06 | 0.000107 | male |
| 20530 | 23122 | 0.2376 | 0.0524 | 5.88E-06 | 0.000107 | male |
| 20530 | 23121 | 0.2353 | 0.0523 | 6.84E-06 | 0.000121 | male |
| 20488 | 23106 | -0.2089 | 0.0468 | 8.17E-06 | 0.000142 | male |
| 20530 | 23109 | -0.2448 | 0.0549 | 8.4E-06 | 0.000143 | male |
| 20530 | 23111 | 0.2718 | 0.0613 | 9.4E-06 | 0.000156 | male |
| 20527 | 23110 | -0.287 | 0.0651 | 1.06E-05 | 0.000173 | male |
| 20497 | 23111 | 0.2462 | 0.0565 | 1.31E-05 | 0.000209 | male |
| 20488 | 23120 | 0.2151 | 0.0497 | 1.51E-05 | 0.000233 | male |
| 20530 | 23125 | 0.2256 | 0.0522 | 1.52E-05 | 0.000233 | male |
| 20530 | 23126 | 0.2229 | 0.0522 | 1.94E-05 | 0.00029 | male |
| 20530 | 23099 | 0.2561 | 0.06 | 1.96E-05 | 0.00029 | male |
| 20498 | 21001 | 0.2592 | 0.0608 | 2.02E-05 | 0.000293 | male |
| 20497 | 23115 | 0.2344 | 0.0552 | 2.16E-05 | 0.000308 | male |
| 20488 | 23128 | 0.2034 | 0.048 | 2.22E-05 | 0.000311 | male |
| 20530 | 23110 | -0.2315 | 0.0548 | 2.44E-05 | 0.000336 | male |
| 20488 | 23124 | 0.2076 | 0.0494 | 2.61E-05 | 0.000353 | male |
| 20488 | 23119 | 0.2014 | 0.0481 | 2.86E-05 | 0.00038 | male |
| 20487 | 23111 | 0.2829 | 0.0679 | 3.07E-05 | 0.000402 | male |
| 20488 | 23127 | 0.1944 | 0.047 | 3.48E-05 | 0.000448 | male |
| 20527 | 21002 | 0.2627 | 0.0638 | 3.87E-05 | 0.00049 | male |
| 20498 | 48 | 0.2601 | 0.0634 | 4.09E-05 | 0.000505 | male |
| 20498 | 23104 | 0.2499 | 0.0609 | 4.11E-05 | 0.000505 | male |
| 20487 | 23115 | 0.2826 | 0.0693 | 4.52E-05 | 0.000547 | male |
| 20498 | 23099 | 0.2494 | 0.0612 | 4.66E-05 | 0.000555 | male |
| 20527 | 23098 | 0.2625 | 0.0647 | 4.95E-05 | 0.000581 | male |
| 20530 | 23127 | 0.2365 | 0.0601 | 8.34E-05 | 0.000959 | male |
| 20488 | 23123 | 0.1897 | 0.0482 | 8.41E-05 | 0.000959 | male |
| 20530 | 49 | 0.2198 | 0.0561 | 8.99E-05 | 0.000998 | male |
| 20487 | 21001 | 0.2598 | 0.0665 | 9.41E-05 | 0.000998 | male |
| 20487 | 23104 | 0.252 | 0.0652 | 0.0001 | 0.000998 | male |
| 20487 | 23112 | 0.2538 | 0.0669 | 0.0001 | 0.000998 | male |
| 20488 | 49 | 0.1871 | 0.049 | 0.0001 | 0.000998 | male |
| 20498 | 23100 | 0.2411 | 0.0623 | 0.0001 | 0.000998 | male |
| 20530 | 23105 | 0.1991 | 0.0513 | 0.0001 | 0.000998 | male |
| 20530 | 23117 | 0.2 | 0.0517 | 0.0001 | 0.000998 | male |
| 20530 | 23118 | 0.1991 | 0.0516 | 0.0001 | 0.000998 | male |
| 20530 | 23123 | 0.2252 | 0.0584 | 0.0001 | 0.000998 | male |
| 20487 | 23116 | 0.2501 | 0.0675 | 0.0002 | 0.001834 | male |
| 20487 | 48 | 0.2449 | 0.0665 | 0.0002 | 0.001834 | male |
| 20498 | 23127 | 0.2313 | 0.0614 | 0.0002 | 0.001834 | male |
| 20527 | 23106 | -0.2426 | 0.0651 | 0.0002 | 0.001834 | male |
| 20530 | 23106 | -0.1971 | 0.0528 | 0.0002 | 0.001834 | male |
| 20530 | 23119 | 0.2185 | 0.0585 | 0.0002 | 0.001834 | male |
| 20488 | 21002 | 0.1791 | 0.0482 | 0.0002 | 0.001834 | male |
| 20487 | 23099 | 0.2437 | 0.0675 | 0.0003 | 0.002494 | male |
| 20488 | 23098 | 0.1755 | 0.0482 | 0.0003 | 0.002494 | male |
| 20491 | 23111 | -0.3448 | 0.0958 | 0.0003 | 0.002494 | male |
| 20491 | 23115 | -0.3427 | 0.0945 | 0.0003 | 0.002494 | male |
| 20497 | 23099 | 0.1884 | 0.0517 | 0.0003 | 0.002494 | male |
| 20498 | 23128 | 0.2274 | 0.0624 | 0.0003 | 0.002494 | male |
| 20527 | 49 | 0.2547 | 0.0698 | 0.0003 | 0.002494 | male |
| 20530 | 23102 | 0.1819 | 0.0504 | 0.0003 | 0.002494 | male |
| 20530 | 23114 | 0.183 | 0.0511 | 0.0003 | 0.002494 | male |
| 20497 | 23112 | 0.1921 | 0.0547 | 0.0004 | 0.003224 | male |
| 20530 | 23101 | 0.1773 | 0.0505 | 0.0004 | 0.003224 | male |
| 20530 | 23113 | 0.1814 | 0.051 | 0.0004 | 0.003224 | male |
| 20498 | 23119 | 0.2045 | 0.0592 | 0.0005 | 0.00395 | male |
| 20498 | 23120 | 0.2166 | 0.0619 | 0.0005 | 0.00395 | male |
| 20498 | 23124 | 0.2133 | 0.0621 | 0.0006 | 0.004694 | male |
| 20491 | 21001 | -0.3023 | 0.0897 | 0.0008 | 0.00608 | male |
| 20497 | 23116 | 0.1809 | 0.0539 | 0.0008 | 0.00608 | male |
| 20497 | 23127 | 0.1702 | 0.051 | 0.0008 | 0.00608 | male |
| 20487 | 23100 | 0.2174 | 0.0662 | 0.001 | 0.007458 | male |
| 20491 | 23104 | -0.2918 | 0.0884 | 0.001 | 0.007458 | male |
| 20487 | 23127 | 0.2213 | 0.0678 | 0.0011 | 0.008128 | male |
| 20522 | 23129 | 0.2232 | 0.069 | 0.0012 | 0.008705 | male |
| 20522 | 23130 | 0.2229 | 0.0689 | 0.0012 | 0.008705 | male |
| 20522 | 23101 | 0.2156 | 0.0669 | 0.0013 | 0.009263 | male |
| 20522 | 23114 | 0.2064 | 0.0644 | 0.0013 | 0.009263 | male |
| 20522 | 20015 | 0.219 | 0.0683 | 0.0014 | 0.009887 | male |
| 20498 | 23110 | -0.2002 | 0.0631 | 0.0015 | 0.010319 | male |
| 20522 | 23102 | 0.2119 | 0.0666 | 0.0015 | 0.010319 | male |
| 20522 | 23113 | 0.2039 | 0.0643 | 0.0015 | 0.010319 | male |
| 20491 | 23112 | -0.2818 | 0.0893 | 0.0016 | 0.01082 | male |
| 20498 | 23123 | 0.1886 | 0.0598 | 0.0016 | 0.01082 | male |
| 20491 | 23116 | -0.276 | 0.0878 | 0.0017 | 0.0114 | male |
| 20522 | 23105 | 0.2041 | 0.0657 | 0.0019 | 0.012531 | male |
| 20522 | 23117 | 0.1989 | 0.064 | 0.0019 | 0.012531 | male |
| 20491 | 23099 | -0.2656 | 0.0859 | 0.002 | 0.012976 | male |
| 20530 | 23130 | 0.1553 | 0.0503 | 0.002 | 0.012976 | male |
| 20488 | 23121 | 0.1441 | 0.0468 | 0.0021 | 0.013195 | male |
| 20522 | 23118 | 0.1975 | 0.0641 | 0.0021 | 0.013195 | male |
| 20527 | 23122 | 0.1965 | 0.0639 | 0.0021 | 0.013195 | male |
| 20530 | 23129 | 0.1548 | 0.0504 | 0.0021 | 0.013195 | male |
| 20525 | 23115 | -0.3575 | 0.117 | 0.0022 | 0.013716 | male |
| 20522 | 50 | 0.2011 | 0.066 | 0.0023 | 0.014118 | male |
| 20527 | 23121 | 0.1945 | 0.0638 | 0.0023 | 0.014118 | male |
| 20488 | 23122 | 0.1423 | 0.0469 | 0.0024 | 0.01462 | male |
| 20525 | 21001 | -0.3073 | 0.1019 | 0.0026 | 0.015718 | male |
| 20488 | 23125 | 0.1398 | 0.0468 | 0.0028 | 0.0168 | male |
| 20487 | 23119 | 0.1995 | 0.067 | 0.0029 | 0.01727 | male |
| 20498 | 21002 | 0.1787 | 0.0601 | 0.003 | 0.017733 | male |
| 20487 | 23128 | 0.1937 | 0.0658 | 0.0032 | 0.018504 | male |
| 20491 | 50 | 0.2391 | 0.0811 | 0.0032 | 0.018504 | male |
| 20525 | 23104 | -0.3009 | 0.1019 | 0.0032 | 0.018504 | male |
| 20488 | 23126 | 0.138 | 0.0469 | 0.0033 | 0.018945 | male |
| 20487 | 23123 | 0.1969 | 0.0673 | 0.0034 | 0.01938 | male |
| 20498 | 23109 | -0.1805 | 0.0619 | 0.0035 | 0.019809 | male |
| 20525 | 23111 | -0.3359 | 0.116 | 0.0038 | 0.021355 | male |
| 20525 | 23116 | -0.3147 | 0.1095 | 0.004 | 0.022322 | male |
| 20491 | 23119 | -0.2354 | 0.0819 | 0.0041 | 0.02241 | male |
| 20491 | 23127 | -0.2423 | 0.0845 | 0.0041 | 0.02241 | male |
| 20497 | 48 | 0.1556 | 0.0543 | 0.0041 | 0.02241 | male |
| 20487 | 23109 | -0.1697 | 0.0592 | 0.0042 | 0.0228 | male |
| 20491 | 23110 | 0.2449 | 0.0862 | 0.0045 | 0.024101 | male |
| 20497 | 21001 | 0.1466 | 0.0516 | 0.0045 | 0.024101 | male |
| 20497 | 23100 | 0.1469 | 0.0518 | 0.0046 | 0.02415 | male |
| 20498 | 23098 | 0.1725 | 0.0609 | 0.0046 | 0.02415 | male |
| 20527 | 23117 | 0.1828 | 0.0645 | 0.0046 | 0.02415 | male |
| 20527 | 23118 | 0.1826 | 0.0646 | 0.0047 | 0.024514 | male |
| 20522 | 23126 | 0.1863 | 0.0661 | 0.0048 | 0.024873 | male |
| 20526 | 48 | 0.2519 | 0.0896 | 0.0049 | 0.025227 | male |
| 20491 | 20015 | 0.2279 | 0.0812 | 0.005 | 0.025577 | male |
| 20488 | 23108 | -0.1296 | 0.0464 | 0.0052 | 0.026431 | male |
| 20525 | 23112 | -0.305 | 0.1093 | 0.0053 | 0.026765 | male |
| 20488 | 23117 | 0.133 | 0.0479 | 0.0054 | 0.026765 | male |
| 20497 | 23104 | 0.1447 | 0.052 | 0.0054 | 0.026765 | male |
| 20527 | 50 | -0.1531 | 0.055 | 0.0054 | 0.026765 | male |
| 20487 | 23110 | -0.172 | 0.0619 | 0.0055 | 0.026926 | male |
| 20527 | 23126 | 0.1793 | 0.0646 | 0.0055 | 0.026926 | male |
| 20528 | 48 | 0.2724 | 0.0988 | 0.0058 | 0.028222 | male |
| 20497 | 23119 | 0.14 | 0.0508 | 0.0059 | 0.028535 | male |
| 20527 | 23125 | 0.1778 | 0.0649 | 0.0061 | 0.029324 | male |
| 20522 | 23125 | 0.1801 | 0.066 | 0.0064 | 0.030582 | male |
| 20487 | 50 | -0.1609 | 0.0592 | 0.0065 | 0.030692 | male |
| 20494 | 23115 | 0.3276 | 0.1204 | 0.0065 | 0.030692 | male |
| 20488 | 23118 | 0.1303 | 0.0479 | 0.0066 | 0.0308 | male |
| 20497 | 50 | -0.127 | 0.0468 | 0.0066 | 0.0308 | male |
| 20522 | 23122 | 0.1781 | 0.0657 | 0.0067 | 0.031085 | male |
| 20487 | 23124 | 0.1766 | 0.0652 | 0.0068 | 0.031366 | male |
| 20487 | 23120 | 0.1761 | 0.0653 | 0.007 | 0.03192 | male |
| 20522 | 23121 | 0.1777 | 0.0659 | 0.007 | 0.03192 | male |
| 20525 | 23110 | 0.2611 | 0.0978 | 0.0076 | 0.034459 | male |
| 20497 | 23128 | 0.1347 | 0.051 | 0.0083 | 0.03742 | male |
| 20491 | 23123 | -0.2115 | 0.0803 | 0.0084 | 0.037658 | male |
| 20494 | 23099 | 0.3192 | 0.1212 | 0.0085 | 0.037894 | male |
| 20494 | 23111 | 0.3107 | 0.1184 | 0.0087 | 0.03857 | male |
| 20488 | 50 | -0.1189 | 0.0456 | 0.0091 | 0.04012 | male |
| 20525 | 23099 | -0.2774 | 0.1075 | 0.0099 | 0.043408 | male |
| 20494 | 23127 | 0.3068 | 0.1196 | 0.0103 | 0.044915 | male |
| 20491 | 23100 | -0.2105 | 0.0823 | 0.0106 | 0.045972 | male |
| 20528 | 23104 | 0.2408 | 0.0944 | 0.0107 | 0.046155 | male |
| 20526 | 21001 | 0.2113 | 0.0829 | 0.0108 | 0.046335 | male |
| 20489 | 50 | 0.0971 | 0.0382 | 0.011 | 0.046691 | male |
| 20525 | 48 | -0.2593 | 0.102 | 0.011 | 0.046691 | male |
| 20528 | 21001 | 0.2386 | 0.094 | 0.0111 | 0.046867 | male |

**Supplemental Table 4:** LCV results among traits related to trauma and social support and anthropometric traits surviving FDR multiple testing correction. **p:** p-value. **zscore:** z-score for partial genetic causality. **gcp.pm:** posterior genetic causality proportion (positive = trait 1 🡪 trait 2). **gcp.pse:** posterior standard error for genetic causality proportion. **h2.zscore.1:** z-score for trait 1 being significantly heritable. **h2.zscore.2:** z-score for trait 2 being significantly heritable.

| **Trait 1** | **Trait 2** | **Sex** | **p** | **FDR q** | **Z.score** | **gcp.pm** | **gcp.pse** | **h2.z.1** | **h2.z.2** | **In text interpretation** |
| --- | --- | --- | --- | --- | --- | --- | --- | --- | --- | --- |
| 20530 | 23114 | female | 3.45E-47 | 4.07E-45 | 27.005 | 0.23 | 0.458 | 8.303 | 52.894 | Witnessed sudden violent death → Leg predicted mass (right) |
| 20530 | 23113 | female | 6.91E-46 | 6.52E-44 | 26.081 | 0.24 | 0.457 | 8.303 | 52.992 | Witnessed sudden violent death → Leg fat-free mass (right) |
| 20530 | 23102 | female | 1.04E-40 | 8.18E-39 | 22.637 | 0.225 | 0.472 | 8.303 | 52.817 | Witnessed sudden violent death → Whole body water mass |
| 20530 | 23101 | female | 1.46E-37 | 9.84E-36 | 20.704 | 0.281 | 0.452 | 8.303 | 52.866 | Witnessed sudden violent death → Whole body fat-free mass |
| 20530 | 23117 | female | 3.65E-31 | 2.15E-29 | 17.095 | 0.052 | 0.493 | 8.303 | 52.906 | Witnessed sudden violent death → Leg fat-free mass (left) |
| 20530 | 23118 | female | 7.51E-26 | 3.94E-24 | 14.363 | 0.035 | 0.496 | 8.303 | 52.736 | Witnessed sudden violent death → Leg predicted mass (left) |
| 20530 | 23129 | female | 2.23E-21 | 1.05E-19 | 12.203 | 0.406 | 0.411 | 8.303 | 52.964 | Witnessed sudden violent death → Trunk fat-free mass |
| 20522 | 50 | male | 2.47E-20 | 4.67E-18 | 11.711 | 0.44 | 0.355 | 9.941 | 41.274 | Been in a confiding relationship as an adult → Standing height |
| 20522 | 20015 | male | 7.82E-20 | 7.39E-18 | 11.477 | 0.623 | 0.335 | 9.941 | 39.101 | Been in a confiding relationship as an adult → Sitting height |
| 20530 | 23130 | female | 3.21E-18 | 1.38E-16 | 10.727 | 0.465 | 0.394 | 8.303 | 53.092 | Witnessed sudden violent death → Trunk predicted mass |
| 20526 | 48 | male | 3.11E-14 | 1.96E-12 | 8.89 | 0.729 | 0.177 | 10.933 | 48.62 | Been in serious accident believed to be life-threatening → Waist circumference |
| 20530 | 23105 | female | 6.45E-12 | 2.54E-10 | 7.809 | -0.094 | 0.547 | 8.303 | 52.449 | Basal metabolic rate → Witnessed sudden violent death |
| 20528 | 23115 | female | 1.94E-11 | 7.04E-10 | 7.583 | 0.776 | 0.15 | 12.563 | 57.615 | Diagnosed with life-threatening illness → Leg fat percentage (left) |
| 20530 | 23126 | female | 2.45E-09 | 8.21E-08 | 6.568 | -0.14 | 0.566 | 8.303 | 52.611 | Arm predicted mass (left) → Witnessed sudden violent death |
| 20526 | 21001 | male | 2.51E-09 | 6.78E-08 | 6.563 | 0.707 | 0.197 | 10.933 | 46.977 | Been in serious accident believed to be life-threatening → Body mass index |
| 20530 | 23125 | female | 2.61E-09 | 8.21E-08 | 6.555 | -0.132 | 0.574 | 8.303 | 52.613 | Arm fat-free mass (left) → Witnessed sudden violent death |
| 20528 | 21001 | female | 4.46E-09 | 1.32E-07 | 6.44 | 0.782 | 0.152 | 12.563 | 51.903 | Diagnosed with life-threatening illness → Body mass index |
| 20491 | 23111 | female | 1.72E-08 | 4.78E-07 | 6.146 | 0.76 | 0.16 | 9.66 | 57.58 | Someone to take to doctor when needed as a child → Leg fat percentage (right) |
| 20528 | 23111 | female | 3.31E-08 | 8.68E-07 | 6.001 | 0.727 | 0.176 | 12.563 | 57.58 | Diagnosed with life-threatening illness → Leg fat percentage (right) |
| 20528 | 23104 | female | 9.27E-08 | 0.0000023 | 5.77 | 0.756 | 0.166 | 12.563 | 51.793 | Diagnosed with life-threatening illness → Body mass index |
| 20530 | 23122 | female | 3.68E-07 | 0.00000868 | 5.455 | -0.118 | 0.532 | 8.303 | 53.087 | Arm predicted mass (right) → Witnessed sudden violent death |
| 20491 | 23115 | female | 4.94E-07 | 0.0000111 | 5.386 | 0.739 | 0.171 | 9.66 | 57.615 | Someone to take to doctor when needed as a child → Leg fat percentage (left) |
| 20491 | 23123 | female | 0.000001 | 0.0000211 | 5.22 | 0.72 | 0.179 | 9.66 | 56.718 | Someone to take to doctor when needed as a child → Arm fat percentage (left) |
| 20491 | 23119 | female | 0.00000103 | 0.0000211 | 5.214 | 0.719 | 0.179 | 9.66 | 56.926 | Someone to take to doctor when needed as a child → Arm fat percentage (right) |
| 20522 | 23129 | male | 0.00000133 | 0.0000314 | 5.154 | -0.011 | 0.075 | 9.941 | 49.075 | Trunk fat-free mass → Been in a confiding relationship as an adult |
| 20528 | 23110 | female | 0.00000244 | 0.000048 | 5.007 | 0.617 | 0.274 | 12.563 | 57.733 | Diagnosed with life-threatening illness → Impedance of arm (left) |
| 20494 | 23127 | male | 0.00000461 | 0.0000968 | 4.852 | 0.724 | 0.183 | 7.146 | 53.856 | Felt irritable or had angry outbursts in past month → Trunk fat percentage |
| 20530 | 23121 | female | 0.0000107 | 0.000202 | 4.642 | -0.143 | 0.528 | 8.303 | 53.307 | Arm fat-free mass (right) → Witnessed sudden violent death |
| 20528 | 23116 | female | 0.0000137 | 0.000249 | 4.581 | 0.726 | 0.182 | 12.563 | 51.712 | Diagnosed with life-threatening illness → Leg fat mass (left) |
| 20491 | 23099 | female | 0.000015 | 0.000262 | 4.556 | 0.658 | 0.192 | 9.66 | 58.612 | Someone to take to doctor when needed as a child → Body fat percentage |
| 20528 | 23099 | female | 0.0000884 | 0.00149 | 4.09 | 0.687 | 0.201 | 12.563 | 58.612 | Diagnosed with life-threatening illness → Body fat percentage |
| 20528 | 23112 | female | 0.000117 | 0.00191 | 4.013 | 0.701 | 0.195 | 12.563 | 51.501 | Diagnosed with life-threatening illness → Leg fat mass (right) |
| 20522 | 23130 | male | 0.000117 | 0.00201 | 4.013 | -0.015 | 0.038 | 9.941 | 49.098 | Trunk predicted mass → Been in a confiding relationship as an adult |
| 20528 | 23119 | female | 0.000136 | 0.00214 | 3.972 | 0.688 | 0.204 | 12.563 | 56.926 | Diagnosed with life-threatening illness → Arm fat percentage (right) |
| 20488 | 48 | male | 0.000169 | 0.00243 | 3.912 | -0.318 | 0.184 | 13.443 | 48.62 | Waist circumference → Physically abused by family as a child |
| 20488 | 21001 | male | 0.00018 | 0.00243 | 3.895 | -0.742 | 0.189 | 13.443 | 46.977 | Body mass index → Physically abused by family as a child |
| 20527 | 23127 | male | 0.000272 | 0.00343 | 3.777 | 0.723 | 0.192 | 7.951 | 53.856 | Been involved in combat or exposed to war-zone → Trunk fat percentage |
| 20528 | 23120 | female | 0.000288 | 0.00431 | 3.761 | 0.696 | 0.2 | 12.563 | 52.653 | Diagnosed with life-threatening illness → Arm fat mass (right) |
| 20491 | 21001 | female | 0.000292 | 0.00431 | 3.757 | 0.696 | 0.202 | 9.66 | 51.903 | Someone to take to doctor when needed as a child → Body mass index |
| 20527 | 49 | male | 0.000293 | 0.00347 | 3.756 | 0.087 | 0.649 | 7.951 | 52.381 | Been involved in combat or exposed to war-zone → Hip circumference |
| 20528 | 23124 | female | 0.000343 | 0.0049 | 3.711 | 0.691 | 0.202 | 12.563 | 52.342 | Diagnosed with life-threatening illness → Arm fat mass (left) at gcp=0.691 |
| 20491 | 23104 | female | 0.000354 | 0.00492 | 3.701 | 0.689 | 0.204 | 9.66 | 51.793 | Someone to take to doctor when needed as a child → Body mass index |
| 20527 | 23099 | male | 0.000395 | 0.00439 | 3.67 | 0.731 | 0.192 | 7.951 | 54.068 | Been involved in combat or exposed to war-zone → Body fat percentage |
| 20488 | 23104 | male | 0.000649 | 0.00681 | 3.523 | -0.739 | 0.196 | 13.443 | 47.192 | Body mass index → Physically abused by family as a child |
| 20491 | 23120 | female | 0.00166 | 0.0223 | 3.236 | 0.643 | 0.218 | 9.66 | 52.653 | Someone to take to doctor when needed as a child → Arm fat mass (right) |
| 20494 | 23099 | male | 0.00176 | 0.0175 | 3.216 | 0.66 | 0.219 | 7.146 | 54.068 | Felt irritable or had angry outbursts in past month → Body fat percentage |
| 20528 | 23123 | female | 0.00177 | 0.0232 | 3.214 | 0.653 | 0.224 | 12.563 | 56.718 | Diagnosed with life-threatening illness → Arm fat percentage (left) |
| 20491 | 23112 | female | 0.0019 | 0.0242 | 3.192 | 0.645 | 0.219 | 9.66 | 51.501 | Someone to take to doctor when needed as a child → Leg fat mass (right) |
| 20528 | 23100 | female | 0.00227 | 0.0283 | 3.134 | 0.654 | 0.218 | 12.563 | 53.926 | Diagnosed with life-threatening illness → Whole body fat mass |
| 20491 | 23124 | female | 0.00266 | 0.0322 | 3.083 | 0.633 | 0.222 | 9.66 | 52.342 | Someone to take to doctor when needed as a child → Arm fat mass (left) |
| 20488 | 23110 | male | 0.00268 | 0.0253 | 3.081 | -0.6 | 0.237 | 13.443 | 52.846 | Impedance of arm (left) → Physically abused by family as a child |
| 20527 | 23115 | male | 0.00281 | 0.0253 | 3.065 | 0.711 | 0.212 | 7.951 | 58.469 | Been involved in combat or exposed to war-zone → Leg fat percentage (left) |
| 20491 | 23116 | female | 0.00323 | 0.0381 | 3.02 | 0.634 | 0.223 | 9.66 | 51.712 | Someone to take to doctor when needed as a child → Leg fat mass (left) |
| 20491 | 23100 | female | 0.0035 | 0.0403 | 2.992 | 0.591 | 0.24 | 9.66 | 53.926 | Someone to take to doctor when needed as a child → Whole body fat mass |
| 20491 | 23127 | female | 0.00399 | 0.0448 | 2.949 | 0.547 | 0.252 | 9.66 | 60.054 | Someone to take to doctor when needed as a child → Trunk fat percentage |

**Supplemental Table 5:** Significant results of polygenic risk score tests between PTSD, traits related to trauma and social support and anthropometric traits.

| **Base** | **Target** | **Sex** | **R^2^** | **FDR Q** |
| --- | --- | --- | --- | --- |
| Someone to take to doctor when needed as a child | Body mass index (BMI) | female | 1.36E-04 | 1.33E-04 |
| Body mass index | Been involved in combat or exposed to war-zone | male | 2.89E-04 | 1.36E-03 |
| Waist circumference | Witnessed sudden violent death | female | 1.68E-04 | 1.36E-03 |
| Hip circumference | Witnessed sudden violent death | female | 1.91E-04 | 1.75E-03 |
| Hip circumference | Diagnosed with life-threatening illness | female | 1.37E-04 | 2.57E-03 |
| Physically abused by family as a child | Waist circumference | male | 1.10E-04 | 4.11E-03 |
| Wais-to-hip ratio (WHR) adjusted by BMI | Someone to take to doctor when needed as a child | female | 1.91E-04 | 4.61E-03 |
| WHR | Someone to take to doctor when needed as a child | female | 1.41E-04 | 1.27E-02 |
| Felt irritable or had angry outbursts in past month | Waist circumference adjusted by BMI | male | 6.08E-05 | 1.63E-02 |
| Body mass index | Physically abused by family as a child | male | 1.53E-04 | 2.29E-02 |
| Waist circumference adjusted by BMI | Someone to take to doctor when needed as a child | female | 6.49E-05 | 3.74E-02 |
| Been involved in combat or exposed to war-zone | Hip adjusted by BMI | male | 5.29E-05 | 4.27E-02 |
| Physically abused by family as a child | BMI | male | 3.57E-05 | 4.45E-02 |
| Leg fat percentage (right) | PTSD | female | 2.53E-03 | 1.61E-08 |
| Leg fat percentage (left) | PTSD | female | 2.01E-03 | 1.58E-07 |
| Body mass index (BMI) | PTSD | female | 1.69E-03 | 1.39E-06 |
| Weight | PTSD | female | 1.83E-03 | 1.44E-06 |
| Hip circumference | PTSD | female | 1.85E-03 | 1.57E-06 |
| PTSD | BMI | female | 1.28E-04 | 4.43E-06 |
| Leg fat mass (left) | PTSD | female | 1.53E-03 | 8.21E-06 |
| Arm fat mass (left) | PTSD | female | 1.34E-03 | 1.63E-05 |
| Leg fat mass (right) | PTSD | female | 1.36E-03 | 2.53E-05 |
| Body fat percentage | PTSD | female | 1.23E-03 | 4.86E-05 |
| Whole body fat mass | PTSD | female | 1.40E-03 | 5.05E-05 |
| Arm fat mass (right) | PTSD | female | 1.17E-03 | 1.14E-04 |
| PTSD | Arm fat percentage (right) | female | 8.98E-05 | 3.77E-04 |
| PTSD | Body mass index (BMI) | female | 7.34E-05 | 5.97E-04 |
| PTSD | Arm fat mass (right) | female | 7.20E-05 | 9.96E-04 |
| Arm fat percentage (right) | PTSD | female | 1.25E-03 | 1.01E-03 |
| PTSD | Leg fat mass (right) | female | 7.21E-05 | 1.16E-03 |
| PTSD | Leg fat percentage (left) | female | 7.82E-05 | 1.17E-03 |
| PTSD | Arm fat mass (left) | female | 7.19E-05 | 1.18E-03 |
| PTSD | Arm fat percentage (left) | female | 7.72E-05 | 1.19E-03 |
| PTSD | Leg fat mass (left) | female | 6.87E-05 | 1.23E-03 |
| Arm fat percentage (left) | PTSD | female | 9.33E-04 | 1.47E-03 |
| Trunk fat mass | PTSD | female | 8.59E-04 | 1.82E-03 |
| PTSD | Whole body fat mass | female | 7.09E-05 | 1.86E-03 |
| PTSD | Leg fat percentage (right) | female | 7.49E-05 | 2.03E-03 |
| PTSD | Body fat percentage | female | 7.56E-05 | 2.44E-03 |
| PTSD | Weight | female | 5.58E-05 | 2.45E-03 |
| Trunk fat percentage | PTSD | female | 7.27E-04 | 4.18E-03 |
| PTSD | Trunk fat mass | female | 6.26E-05 | 4.83E-03 |
| PTSD | Trunk fat percentage | female | 7.14E-05 | 7.73E-03 |
| PTSD | Hip circumference | female | 3.59E-05 | 1.47E-02 |
| PTSD | Impedance of whole body | female | 3.01E-05 | 1.64E-02 |
| BMI | PTSD | female | 8.60E-04 | 1.78E-02 |
| Impedance of arm (left) | PTSD | female | 3.12E-04 | 3.17E-02 |
| PTSD | Impedance of arm (left) | female | 2.28E-05 | 3.69E-02 |

**Supplemental Table 6:** Two-sample MR results (IVW method). **SE:** standard error. **nSNPs:** number of single nucleotide polymorphisms.

| **Exposure** | **Outcome** | **Sex** | **nSNPs** | **Beta** | **se** | **p** | **FDR Q** |
| --- | --- | --- | --- | --- | --- | --- | --- |
| Leg fat percentage right | PTSD | female | 1468 | 0.319 | 0.053 | 3.13E-09 | 6.89E-08 |
| Weight UKB | PTSD | female | 582 | 0.282 | 0.059 | 1.79E-06 | 1.53E-05 |
| BMI UKB | PTSD | female | 1564 | 0.241 | 0.051 | 2.09E-06 | 1.53E-05 |
| Hip circumference UKB | PTSD | female | 1131 | 0.256 | 0.056 | 5.18E-06 | 2.85E-05 |
| Someone to take to doctor when needed as a child | BMI GIANT | female | 5 | -0.594 | 0.121 | 1.09E-05 | 4.80E-05 |
| Whole body fat mass | PTSD | female | 1043 | 0.232 | 0.053 | 1.48E-05 | 4.81E-05 |
| Leg fat mass left | PTSD | female | 1560 | 0.222 | 0.051 | 1.62E-05 | 4.81E-05 |
| Arm fat mass left | PTSD | female | 1596 | 0.218 | 0.051 | 1.75E-05 | 4.81E-05 |
| WHRadjBMI | Someone to take to doctor when needed as a child | female | 685 | -0.032 | 0.008 | 4.03E-05 | 9.85E-05 |
| Arm fat mass right | PTSD | female | 1205 | 0.203 | 0.052 | 1.02E-04 | 2.24E-04 |
| Body fat percentage | PTSD | female | 1670 | 0.202 | 0.052 | 1.22E-04 | 2.44E-04 |
| Arm fat percentage right | PTSD | female | 131 | 0.377 | 0.099 | 1.41E-04 | 2.59E-04 |
| Trunk fat mass | PTSD | female | 142 | 0.326 | 0.091 | 3.58E-04 | 6.06E-04 |
| Physically abused by family as a child | WC | female | 1175 | 0.055 | 0.015 | 5.07E-04 | 7.97E-04 |
| Arm fat percentage left | PTSD | female | 134 | 0.331 | 0.096 | 6.51E-04 | 9.55E-04 |
| HIP GIANT | Witnessed sudden violent death | female | 346 | 0.013 | 0.004 | 8.62E-04 | 1.19E-03 |
| BMI GIANT | PTSD | female | 189 | 0.273 | 0.083 | 1.02E-03 | 1.32E-03 |
| Trunk fat percentage | PTSD | female | 1304 | 0.162 | 0.053 | 2.46E-03 | 3.01E-03 |
| WCadjBMI | Someone to take to doctor when needed as a child | female | 1384 | -0.020 | 0.007 | 6.70E-03 | 7.76E-03 |
| BMI GIANT | Physically abused by family as a child | male | 353 | 0.028 | 0.011 | 8.19E-03 | 9.01E-03 |
| PTSD | Trunk fat percentage | female | 351 | 0.004 | 0.001 | 1.40E-02 | 1.47E-02 |
| Been involved in combat or exposed to war-zone | HIPadjBMI | male | 959 | -0.111 | 0.049 | 2.50E-02 | 2.50E-02 |
